# Comparison of methods for assessing effects of risk factors on disease progression in Mendelian randomization under index event bias

**DOI:** 10.64898/2026.02.26.26347193

**Authors:** Linxuan Zhang, Ixavier Alonzo Higgins, Qile Dai, Apostolos Gkatzionis, Jocelyn Quistrebert, Nasir Bashir, Gopuraja Dharmalingam, Pallav Bhatnagar, Dipender Gill, Yushi Liu, Stephen Burgess

## Abstract

Mendelian randomization has emerged as a transformative approach for inferring causal relationships between risk factors and disease outcomes. However, applying Mendelian randomization to disease progression – a critical step in validating pharmacological targets – is hampered by index event bias. This form of selection bias occurs because analyses of disease progression are necessarily restricted to individuals who have already experienced the disease event. Here, we present a comprehensive evaluation of statistical methods designed to mitigate index event bias, including inverse-probability weighting, Slope-Hunter, and multivariable methods. We compare the performance of these methods in simulations and applied examples. Inverse-probability weighting methods reduce bias, but require individual-level data and will only fully eliminate bias when the disease event model is correctly specified. Slope-Hunter performed poorly in all simulation scenarios, even when its assumptions were fully satisfied. Multivariable methods worked best when including genetic variants that affect the incident disease event. However, if these genetic variants also affect disease progression directly, then the analysis will suffer from pleiotropy. Hence, if the same biological mechanisms affect disease incidence and progression, then multivariable methods will have little utility. But in such a case, analyses of disease progression are less critical, as conclusions reached from analyses of disease incidence are likely to hold for disease progression. Our findings indicate that no single method is a universal solution to provide reliable results for the investigation of disease progression. Instead, we propose a strategic framework for method selection based on data availability and biological context.

## 1 Introduction

Mendelian randomization is an epidemiological technique that can be used to assess whether a proposed risk factor has a causal effect on a disease outcome [1, 2]. The approach uses genetic predictors of the risk factor which, under the instrumental variable assumptions, will associate with a disease outcome if the risk factor has a causal effect on the outcome [3]. An instrumental variable is a variable that affects the risk factor in a specific way, such that:

1. it is associated with the risk factor,
2. it is not associated with the outcome via a confounding pathway, and
3. it has no direct effect on the outcome, only potentially an indirect effect via the risk factor [4, 5].

Genetic variants are plausible instrumental variables for several reasons. Genes have specific biological functions, and hence genetic variants within these genes plausibly affect biological mechanisms via particular biological pathways. Genetic variants are inherited in a random way, depending on choice of sexual partner and natural variability in gamete formation, and hence should only associate with traits that they affect [6]. Genetic associations are protected from many sources of confounding, as an individual’s genotype is fixed at conception (aside from somatic mutations), meaning that it cannot be affected by environmental confounders or lifestyle choices. The fixed nature of the genotype also provides some protection from bias due to reverse causation [7]. However, there are also many reasons why the instrumental variable assumptions may be violated for a specific genetic variant: for instance, the variant may be pleiotropic (that is, it affects multiple traits on different causal pathways) [8], or it may be subject to population stratification (that is, it is associated with latent population structure) [9]. However, empirically speaking, genetic variants have been shown to be no more associated with confounders than would be expected due to chance alone [10, 11], and well-conducted Mendelian randomization investigations typically agree in the direction of their estimate with results from randomized trials [12].

In principle, Mendelian randomization can be used with a time-to-event outcome, such as disease progression. An association between the outcome and a genetic variant that is a valid instrumental variable, which could be tested using Cox proportional hazards regression (or any other suitable method), would be indicative of a causal effect of the risk factor on the outcome [13]. However, the ‘randomization’ on which Mendelian randomization is based occurs for the population as a whole at conception [14], and so Mendelian randomization should be attempted in datasets that are representative of the population at the point of randomization. It is plausible that this representativeness holds for population-based datasets of middle-aged individuals (Figure 1). Additionally, genetic associations with rare diseases estimated in case-control datasets are unaffected by the sampling strategy, and so should be representative of associations estimated in the whole population. However, the only individuals with data on a disease progression outcome are those who have had a disease event. Hence the dataset for a disease progression outcome is typically not representative of the population at the point of randomization [15]. Further, if the risk factor affects the probability of the disease event, then the disease event is a common effect of the genetic variants and risk factor–disease confounders [16]. Conditioning on the disease event would induce collider bias, leading to an association between the genetic instrument and disease progression even in the absence of a causal effect of the risk factor on disease progression [17]. As an alternative explanation for this problem, taking the analogy of Mendelian randomization with a randomized trial, the disease event is a post-randomization covariate, and hence stratification on this variable is inappropriate [18].

**Figure 1.**
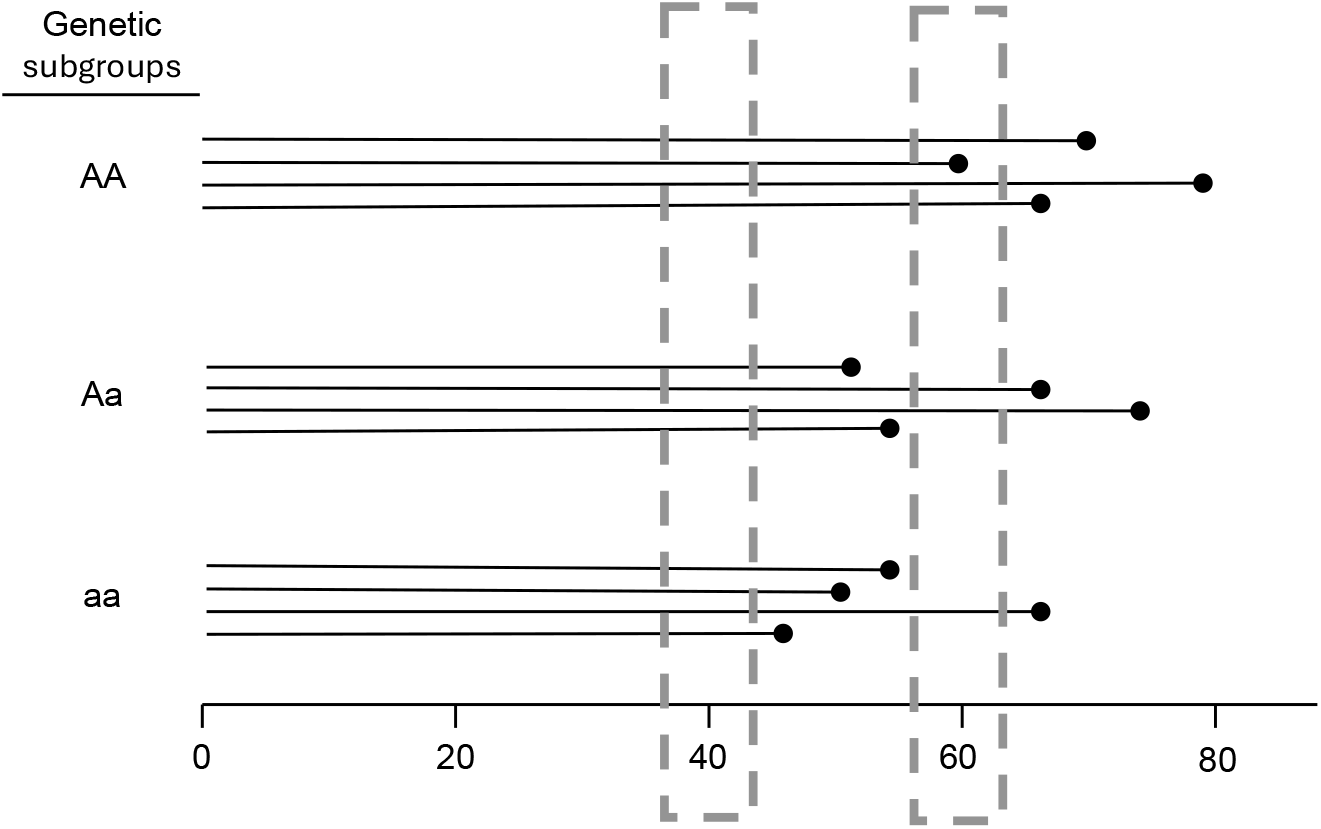
Illustrative diagram of Mendelian randomization with a time-to-event outcome. We assume that the three genetic subgroups are exchangeable at time zero (the point of randomization, which is conception). For many outcomes, if we sample individuals at age 40, then plausibly the groups in our sample are still exchangeable, assuming that almost all individuals survive and are disease free until age 40, or else we have data on health outcomes before recruitment (such as electronic health records). The dataset at age 40 is still representative of the dataset at time zero. However, if we sample individuals at age 60 (or after any selection event), then the groups are no longer exchangeable. The individuals in the genetic subgroup with the worst survival (in this diagram, the aa subgroup) who remain are likely to be the fittest members of this subgroup. Hence, associations estimated in the dataset at age 60 will no longer equal associations estimated in the dataset at time zero.

Several statistical methods have been proposed to deal with this bias, which is a specific example of selection bias known as index event bias [19]. In this work, we compare the performance of five methods: inverse-probability weighting [20], Heckman’s method [21], Slope-Hunter [22], multivariable Mendelian randomization [23], and the corrected weighted bivariate least squares (CWBLS) method [24].

We are particularly interested in the context where the risk factor is a biomarker for a druggable mechanism, and the genetic variants mimic interventions on this mechanism [25]. In such a case, the genetic variants are often taken from a single gene region [26]. If the risk factor is a protein, then this gene region is typically the *cis*-gene region for the protein; that is, the gene region encoding the protein. Mendelian randomization investigations using genetic variants from a single gene region (or small number of gene regions) with strong biological links to the risk factor are known as *cis*-Mendelian randomization investigations [27]. When the risk factor is a proxy for a druggable mechanism, the approach is known as drug target Mendelian randomization [28]. Index event bias is particularly important in this context because the majority of drugs are given to selected groups of patients with a disease to prevent disease progression, not to the whole population.

This manuscript is structured as follows. In Section 2, we provide an introduction to Mendelian randomization and present methods for the analysis of time-to-event data, including those that attempt to account for index event bias. In Section 3, we perform a simulation study to compare the statistical properties of estimates from these methods to analyse synthetic data under index event bias in a range of scenarios. In Section 4, we present examples of Mendelian randomization investigations with disease severity as an outcome, estimated in a selected sample of the population, to assess how the methods perform in a practical example. We conclude by summarizing our findings and with recommendations for applied practice (Section 5).

## 2 Methods

### 2.1 Individual-level data

There are two paradigms under which Mendelian randomization investigations are typically performed [29]. The first paradigm uses individual-level data on a single dataset (known as one-sample Mendelian randomization). The second paradigm uses summarized data on two datasets: one in which we estimate genetic associations with the risk factor (which we refer to as an exposure), and the other in which we estimate genetic associations with the outcome. This categorization is not exhaustive; it is possible that an investigation uses individual-level data on two datasets, or summarized data taken from a single dataset.

Mendelian randomization investigations for a time-to-event outcome can be performed using either paradigm. With individual-participant data, a two-stage method can be employed [13]. In the first stage, we regress the exposure on the genetic variants using linear regression. In the second stage, we regress the outcome on fitted values of the exposure from the first stage. With a time-to-event outcome, we here use Cox proportional hazards regression in the second stage, although Aalen additive hazards regression or another method could also be used [30].

Estimates from Cox regression (similarly to those from logistic regression) are non-collapsible, meaning that a causal effect on the hazard ratio scale typically differs when averaged across the population even if it is constant for all individuals in the population [31]. However, as the primary goal of Mendelian randomization is to detect the existence of a causal effect, this does not overly concern us, as we still have a valid test of the causal null hypothesis. Given that Cox regression is more familiar to epidemiologists and the multiplicative hazards assumption is more reasonable [32], we use Cox regression in this work.

We assume throughout this manuscript that all relationships between variables are linear on the relevant scale. While this is a strong assumption, if genetic associations are small in magnitude (as is typical in practice), then it is a reasonable approximation, as all continuous functions are linear to a first-order approximation.

A summary of the methods considered in this paper is provided as Table 1.

**Table 1:** Summary of index event bias methods considered in this paper.

| Method | Data format | Key assumptions and limitations |
| --- | --- | --- |
| Inverse-probability weighting | Individual | Requires a prediction model for the probability of selection (typically this is the probability of having an incident disease event). Full correction for index event bias is only achieved when this model is fully and correctly specified. |
| Heckman's | Individual | Requires an instrument for the incident disease event. Can only be used for a continuous outcome or a binary outcome. |
| Slope-Hunter | Summarized | Assumes a homogeneous form of index event bias. Uses GWAS data to estimate the degree of bias, under the further assumption that some variants affect the incident disease event and not disease progression, and other variants affect disease progression and not the incident disease event. |
| Multivariable IVW | Summarized | Requires genetic variants that affect the incident disease event in a specific way (that is, they are instruments for the disease event). Fits a multivariable model with disease progression as the outcome, and the risk factor and disease event as the two exposures. |
| CWBLS | Summarized | Same as the multivariable IVW method, but designed to be more robust to weak instruments. |

### 2.2 Inverse-probability weighting

A common method to account for selection bias is inverse-probability weighting. Suppose that we have a statistical model for the probability of selection into the dataset (known as the propensity score) that holds for each individual in the population. If individuals with a particular set of characteristics have a 25% chance of being selected into the dataset, then we would expect to see only 25% of these individuals in our dataset. Hence, if we upweight each of these selected individuals by 4 (= 1/0.25), then we would obtain as many of these individuals as appear in the population. By weighting on the reciprocal of the probability of selection, we obtain a weighted pseudo-population that is representative of the true population as if everyone were selected [20].

In Mendelian randomization, the dataset in which we estimate genetic associations with disease progression typically consists of individuals who had the disease outcome. If we calculate the probability of getting the disease, and then inverse-weight on these probabilities, our dataset should be similar to the underlying population, and hence genetic associations with disease progression in this weighted population should be unbiased [16]. A recent methodological investigation emphasized the importance of modelling interaction terms in such analyses [33].

The selection model is not required to have a causal interpretation. However, the degree of bias reduction will depend on the predictive accuracy of this model. A further practical limitation of the inverse-probability weighting method is its reliance on individual-level data. For practical reasons, Mendelian randomization investigations are often conducted using summarized data, and investigators do not have access to individual-level data.

### 2.3 Heckman’s sample selection method

Heckman’s sample selection method uses an instrument for the selection event to correct for selection bias [34]. In the context of index event bias, the selection event is the incident disease, and hence the model requires an instrumental variable for the disease event. The method combines a regression model with a selection model. These models are fitted jointly, allowing for correlation in the error terms between the models. The selection model is a probit model that includes the instrument for the selection event and any other relevant covariates. The regression model excludes the instrument for the selection event [35]. These models can be fitted using maximum likelihood estimation.

We implement Heckman’s method using the `sampleSelection` package for the R software environment under its default options [36]. This package enables estimation for a continuous outcome or a binary outcome, but not for a time-to-event outcome. Similarly to the inverse-probability weighting method, this method uses individual-level data.

### 2.4 Summarized data

By summarized data, we mean the beta-coefficients and standard errors representing the associations of variants with the risk factor and outcome. Indexing genetic variants by *j*, the inverse-variance weighted (IVW) method regresses genetic associations with the outcome 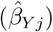 on genetic associations with the risk factor 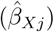 using the reciprocals of the variances of genetic associations with the outcome as weights 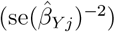 [37]. This regression model does not have an intercept term, as by the instrumental variable assumptions, a genetic instrument having zero association with the risk factor would have zero association with the outcome:

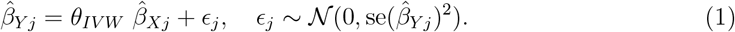

If the same regression method is used for the outcome in the second stage of a two-stage method and in the calculation of summarized associations with the outcome, then the two-stage and IVW methods are related. With a single genetic variant, these two methods give the same answer: the ratio of the coefficients: 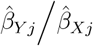. With multiple genetic variants, they give the same answer if the genetic variants are perfectly uncorrelated [38].

Several methods have been proposed for the analysis of summarized data with multiple independent genetic variants that are robust to some of the variants being invalid instruments [39]. The underlying concept is that each valid instrument (under linearity and homogeneity) provides an estimate of the same quantity: the effect of the risk factor on the outcome [40]. With sufficient data, we would expect similar estimates for all genetic variants that are valid instruments, and different estimates for genetic variants that are not valid instruments. If we assume that most genetic variants are valid instruments, then we can develop an estimation strategy that only takes information from genetic variants with similar estimates; under our assumption, these variants are the valid instruments. Alternatively, we can assume that the invalid genetic variants are invalid in a specific way that allows the invalidity to be modelled.

### 2.5 Summarized data with index event bias

Under assumptions of linearity and homogeneity, index event bias in the genetic associations with the outcome should be linear in the genetic associations with the exposure [19]. We further assume that the proportionality constant is the same for all variants. That is, the true genetic association with the outcome as estimated without selection bias is equal to the genetic association with the outcome as estimated in the selected dataset plus the genetic association with the exposure multiplied by a constant. This constant can be estimated using genome-wide association data.

If we write 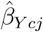 for the genetic association with the outcome as estimated in the selected dataset, 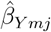 for the genetic association with the outcome as estimated without selection bias, and 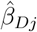 for the genetic associations with disease risk, then

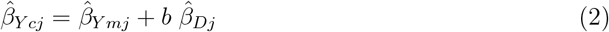

where *b* is a constant to be estimated, which we refer to as the collider bias constant.

Under this index event bias model, the ratio of the coefficients should 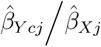 be similar for all genetic variants that are valid instruments. This allows the use of robust methods for Mendelian randomization in the case of index event bias.

### 2.6 Slope-Hunter

The Slope-Hunter method uses model-based clustering to estimate the collider bias constant *b* from the biased genetic associations with the disease progression outcome 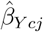, the genetic associations with disease risk 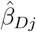, and the genetic associations with the risk factor 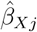 [22]. Slope-Hunter first identifies genetic variants strongly associated with disease risk from a genome-wide association study (GWAS) for disease incidence. It then fits a bivariate normal mixture model to cluster variants into those affecting incidence only (*β*_*Dj*_ ≠ 0, *β*_*Y mj*_ = 0) and those affecting both incidence and progression (*β*_*Dj*_ ≠ 0, *β*_*Y mj*_ ≠ 0). As the true variant-outcome associations *β*_*Y mj*_ are unknown and the estimates 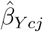 are potentially biased, the clustering algorithm identifies the largest group of genetic variants with similar ratios 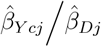 and assumes these are the incidence-only variants; this is called the zero-modal residual assumption (ZEMRA) and is similar to the plurality assumption made by the mode-based Mendelian randomization method [41]. Slope-Hunter then fits the linear regression model 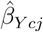 on 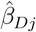 using the incidence-only SNPs and estimates the collider bias constant *b* as the slope of the regression line. The estimate 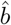 can then be used to correct the summary statistics for all genetic associations with the progression outcome.

Implementation of the Slope-Hunter method requires specifying a tuning parameter that represents the p-value threshold for identifying variants associated with disease incidence. The algorithm can be sensitive to the value of this threshold.

### 2.7 Multivariable Mendelian randomization

Multivariable Mendelian randomization is an extension of standard (univariable) Mendelian randomization when there are multiple exposures of interest [23]. If we have genetic variants that affect a set of exposures in a specific way, such that:

1. each variant is associated with at least one of the exposures,
2. no variant is associated with the outcome via a confounding pathway, and
3. no variant has a direct effect on the outcome, only potentially an indirect effect via one or more exposures.

Additionally we require that each exposure is associated with at least one genetic variant, and there is not perfect collinearity in the genetic associations with the exposures.

Multivariable Mendelian randomization can be implemented by weighted regression of the genetic associations with the outcome on the genetic associations with the exposures. For two exposures, we can form the regression equation:

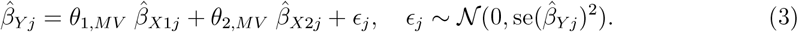

As this regression equation is an extension of that used in the IVW method for the univariable case, we refer to this as the multivariable IVW method.

Suppose that the outcome *Y* is a time-to-event outcome, and *D* is the disease event. Assuming that *Y* and *D* are both affected by the exposure, the coefficient for the exposure from a multivariable IVW model where the exposures are the risk factor and the disease event estimates the effect of the risk factor on the outcome [42]. Adjustment for genetically-predicted risk of the disease event corrects for the index event bias:

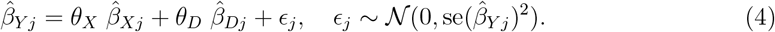

A limitation of this approach is that we need at least as many genetic variants in the model as there are parameters that we estimate (in this case, we need at least two genetic variants). If the genetic variants do not strongly predict the exposures, then we have weak instruments [43]. In particular, the model requires that the genetic associations with the exposures are not collinear, meaning that we do not only need strong instruments, but conditionally strong instruments [44]. Weak instruments lead to bias in univariable Mendelian randomization, but in a two-sample setting, this bias is typically in the direction of the null, and hence does not lead to false positive findings. However, in the context of multivariable Mendelian randomization, weak instrument bias is potentially more serious, as it is more likely (due to trying to predict more exposures) and it can lead to false positive findings [45].

A further limitation of this approach is that we need to find genetic predictors of the disease event that plausibly do not directly affect disease progression.

### 2.8 Corrected weighted bivariate least squares

The corrected weighted bivariate least squares (CWBLS) method is an extension of multivariable Mendelian randomization to account for weak instrument bias [24]. It estimates the amount of bias due to uncertainty in the genetic associations with the exposures, and subtracts this from the estimates. With a single exposure, it is equivalent to the debiased IVW method [46], which has also been expanded to the multivariable setting [47].

## 3 Simulation study

We conduct a simulation study to assess the impact of collider bias on Mendelian randomization estimates, and the performance of methods for dealing with index event bias. Our list of methods is not complete: any multivariable Mendelian randomization method for summarized data could be used (for instance, Donovan et al. [48] used the MR-Horse method [49] as a convenient pleiotropy-robust multivariable method). However, these methods are chosen for comparison because they make different assumptions and use data in different ways:

- the inverse-probability weighting method uses individual-level data, and addresses index event bias by modelling the probability of an incident disease event;
- Heckman’s method uses an instrument for the disease event to account for the selection event;
- the Slope-Hunter method uses summarized genome-wide association data to estimate the collider bias constant, and calculates a corrected slope coefficient using genetic predictors of the risk factor;
- the multivariable inverse-variance weighted method uses summarized data generated from our simulated individual-level data on a limited number of variants, and uses genetic associations with the disease event to correct for index event bias;
- the CWBLS method does the same, but it is designed to account for weak instrument bias.

We additionally perform analyses using the ratio method on the whole population (assuming that everyone has a time-to-event outcome) and only on individuals who have a disease event. The ratio method is equivalent to the two-stage method with a single instrumental variable, which in our simulation study is taken as an unweighted genetic score calculated by summing up the instrumental variants.

### 3.1 Data generating model and scenarios

We simulate data on independent genetic variants, a risk factor, an index disease event, and a time-to-event outcome for 100 000 individuals (one-sample setting) indexed by *i* and genetic variants indexed by *j*. A diagram indicating the data generating mechanism is provided as Figure 2. The genetic variants (*G*_*j*_) are assumed to have independent standard normal distributions. The risk factor (*X*) is modelled as a linear function of the first *J*_1_ genetic variants (which we refer to as the instrumental variants), two independent standard normally distributed confounders (*U*_1_ and *U*_2_), and an independent standard normally distributed error term (*ϵ*_*X*_):

**Figure 2.**
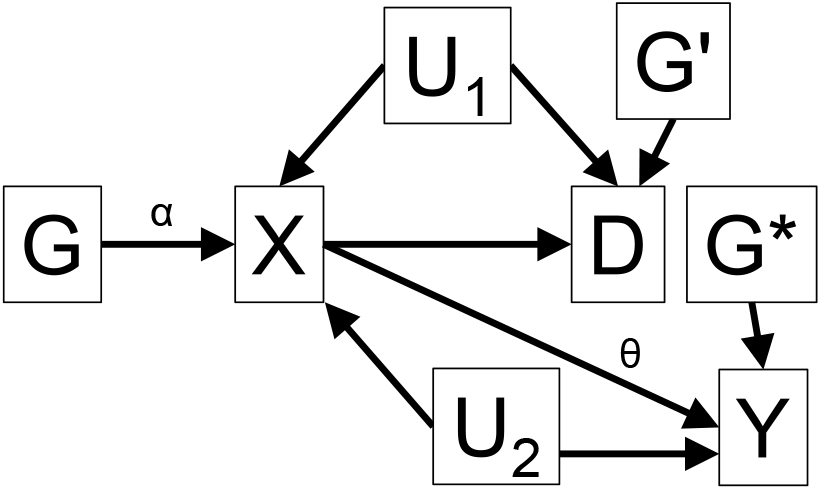
Illustrative diagram of data generating mechanism for simulation study.

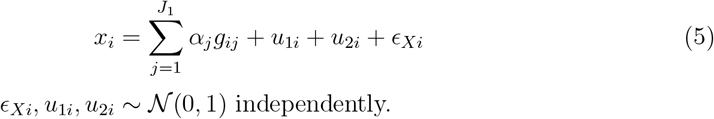

The disease event (*D*) is modelled as a binary event whose probability is a logistic-linear function of the risk factor, one of the confounders, and a separate set of *J*_2_ genetic variants 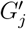 that affect the disease event probability directly:

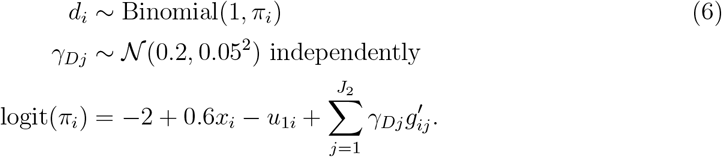

The time-to-event outcome (*Y*) is modelled as an exponential variable whose mean is a log-linear function of the risk factor (with effect parameter *θ*), the other confounder, and a third separate set of *J*_3_ genetic variants 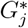 that affect the outcome directly:

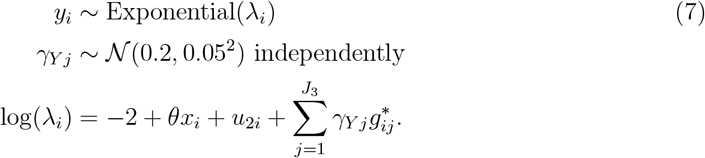

For simplicity, in the individual-level data analyses, we construct an unweighted genetic score based on the *J*_1_ instrumental variants, and use this variable as the sole instrument. In the inverse-probability weighting method, the propensity score models the logit-transformed probability of the disease event as a function of the risk factor, this genetic score, and their interaction. We note that this model is not fully specified, as the confounder *U*_1_ is assumed to be unknown and the model does not include the additional variants 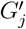. In Heckman’s method, we treat the time-to-event outcome as a continuous variable, and assess the association between the genetic score and this outcome in individuals with an incident disease event. In the selection model, we fit the selection event on the genetic score and the 10 additional variants that affect the disease event directly. For the summarized data methods, we calculate the genetic associations with the risk factor (using linear regression), the disease event (using logistic regression), and the time-to-event outcome (using Cox regression); the data used by the methods are the beta-coefficients and standard errors from separate regressions on each genetic variant in turn. The genetic associations are estimated in the same individuals. Hence this is a one-sample investigation, although given the genetic associations are strong, any bias due to the sample overlap should be minimal.

For the Slope-Hunter method, we simulate association data for 1000 additional variants, whose associations with the disease event and time-to-event outcome are drawn from normal distributions with mean zero. This is to represent genome-wide association data, which is used by the algorithm to estimate the amount of collider bias. The genome-wide association data consists of the *J*_1_ instrumental variants, the *J*_2_ variants that affect the disease event directly, the *J*_3_ variants that affect the time-to-event outcome directly, and the 1000 additional null variants. For the multivariable methods, we run analyses two ways. First, we run analyses based on the *J*_1_ instrumental variants only (“IVs only”). Second, we run analyses based on the *J*_1_ instrumental variants and the *J*_2_ variants that affect the disease event directly (“including disease”).

Illustrative code is available in the Supplementary Material, and full simulation code can be found at https://github.com/sb452/mrprog. All methods were performed using their default parameter settings. The Slope-Hunter method was performed using 100 bootstrap samples per iteration. For the inverse-probability weighting method, we report the robust standard errors.

We consider two main settings: one in which the effect of the risk factor on the time-to-event outcome is positive (*θ* = +0.2), and one in which it is null (*θ* = 0). This provides insight into the performance of the methods when there is a causal effect of the risk factor, and when there is not. We are interested in the median estimate across simulated datasets, the median standard error of estimates, and the proportion of datasets that include zero in their 95% confidence interval. With a positive causal effect, this is the empirical power of the analysis; with a null causal effect, it is the empirical Type 1 error rate (also called the coverage). We describe estimates with a p-value below 0.05 as ‘significant’.

For each setting, we consider six scenarios: three in which we vary the instrument strength, and three in which we vary the number of instruments. In the first case, we consider *J*_1_ = 10 genetic instrumental variants and draw the genetic effects on the risk factor *α*_*j*_ from a normal distribution with standard deviation 0.02 and mean of 0.25 (stronger instruments), 0.15 (moderate instruments), and 0.08 (weaker instruments). In the second case, we use the moderate instruments setting, and consider 5, 10, and 20 genetic instruments. We note that these descriptors are relative: the instruments in the ‘weaker instruments’ case are still reasonably strong, with an average F statistic for the genetic score of 2500. The average proportion of variance in the risk factor estimated was around 17% for the stronger instruments, 7% for the moderate instruments, and 2% for the weaker instruments. Initially, we consider *J*_2_ = 10 variants that directly affect the disease event, and *J*_3_ = 10 variants that directly affect the time-to-event outcome. For each set of parameter values, we simulate 1000 datasets. A diagram illustrating the data generating model is provided as Figure 2.

We note that this simulation study is deliberately intended to be a ‘friendly’ example – that is, one in which all the model assumptions are satisfied, and the methods should all perform well. The aim of this simulation is not to determine how the methods might behave in a realistic setting, but to explore how they perform relative to each other.

We also vary the data generating model to consider different conditions as secondary scenarios:

- We vary the direction of confounding effects on the disease event and the time-to-event outcome.
- We use a common confounder for the risk factor with both the disease event and the time-to-event outcome, and we introduce a separate confounder of the disease event and time-to-event outcome.
- We add a gene–confounder interaction term in the effect of the genetic variants on the exposure.
- We vary the number of additional variants that directly affect the disease event and the time-to-event outcome in the Slope-Hunter and multivariable methods.
- We vary the number of additional variants that directly affect the disease event in Heckman’s method, using a binary outcome.
- We use variants that affect both the disease event and the time-to-event outcome in the Slope-Hunter and multivariable methods.

Full details of all simulation models are provided in the Supplementary Material.

### 3.2 Simulation results

A summary of results for the moderate strength scenario with 10 instruments is shown in Table 2.

**Table 2:** Summary of simulation results for a single base scenario: moderate strength, 10 instruments.

| Method | Positive effect |  | Null effect |  |
| --- | --- | --- | --- | --- |
|  | Median | Power | Median | Coverage |
| Ratio method, full sample | 0.103 | 100.0 | 0.000 | 4.9 |
| Ratio method, selected sample | 0.072 | 98.6 | -0.028 | 41.6 |
| Inverse-probability weighting | 0.091 | 99.4 | -0.016 | 12.7 |
| Slope-Hunter | 0.085 | 71.5 | 0.000 | 52.7 |
| Multivariable IVW (IVs only) | 0.107 | 7.7 | 0.004 | 3.5 |
| Corrected weighted bivariate (IVs) | 0.036 | 3.2 | -0.052 | 2.2 |
| Multivariable IVW (including disease) | 0.105 | 100.0 | 0.000 | 3.7 |
| Corrected weighted bivariate (disease) | 0.106 | 100.0 | 0.000 | 7.5 |

The full sample analysis represents a gold-standard analysis, but typically an unachievable one in a realistic setting, as we assume that we have full outcome data on all individuals, not just those with a disease event. With a positive effect, the median estimate from the full sample analysis is 0.103. This is less than *θ* = 0.2 due to non-collapsibility; however, it is clearly a positive effect, and it is detected in 100% of simulated datasets. With a null effect, the median estimate is almost exactly zero, and a significant effect is detected in 4.9% of datasets, in line with the expected 5% false positive rate of a valid 95% confidence interval.

The selected sample (case-only) analysis represents a naive analysis that does not account for index event bias. With a positive effect, estimates are large enough that the index event bias does not change our findings. However, with a null effect, the median estimate is less than zero, and a significant effect is detected in 41.6% of datasets under the null, far more than the expected 5% false positive rate of a valid 95% confidence interval.

The inverse-probability weighting method is less biased than the selected sample analysis, indicating that the method can reduce index event bias. It also maintains high power, with a positive effect detected in 99.4% of simulated datasets. However, we still see some bias and Type 1 error inflation under the null, with a significant effect detected in 12.7% of datasets under the null. The reason why we see Type 1 error inflation is that the selection model is not perfectly specified, as the probability of having a disease event is a function of the confounder (which is assumed to be unmeasured). If this model were correctly specified, we would expect to have perfectly unbiased estimates with nominal Type 1 error rates.

Results from Heckman’s method are not provided in Table 2 as they were substantially different from other methods. The method estimated associations between the genetic score and the outcome using a linear model, and hence the estimates are not comparable to those from other methods. Additionally, the method occasionally gave infinite confidence intervals. When changing the time-to-event outcome to have a normal distribution, estimates were more reasonable. We do not consider Heckman’s method further in the main simulations; however, we perform an additional scenario with a binary outcome variable to assess its performance.

The Slope-Hunter method has good power to detect a causal effect, and the median estimate is unbiased under the null. However, Type 1 error inflation is greater in magnitude for the Slope-Hunter method compared to the naive (selected sample) method, with a significant effect detected by the Slope-Hunter method in 52.7% of datasets under the null.

When only including instrumental variants, the multivariable IVW method gives close to median unbiased estimates under the null and maintains nominal Type 1 error rates. However, power to detect a positive effect is much lower, at 7.7%. The CWBLS method is similar, but has even lower power (3.2%). When including genetic predictors of the disease event, the multivariable IVW and CWBLS methods have 100% power. Coverage for the multivariable IVW method is in line with nominal levels (3.7%), whereas coverage for the CWBLS method is slightly above nominal levels (7.5%). For reference, for a true 5% test and 1000 replications, we would expect 95% of our observations for the empirical coverage to lie between 3.7% and 6.4%.

Results varying the strength of instruments are provided in Table 3, and results varying the number of instruments are provided in Table 4. Generally speaking, findings consistently show that the inverse-probability weighting and Slope-Hunter methods have reasonable power to detect a positive causal effect, but both methods have inflated Type 1 error rates under the null, particularly for Slope-Hunter. When only including instrumental variants, the multivariable IVW and CWBLS methods have controlled Type 1 error rates under the null, but much lower power to detect a positive causal effect. When including genetic predictors of the disease event, the multivariable IVW and CWBLS methods have high power. Coverage for the multivariable IVW method remains in line with nominal levels in all cases (between 3.7% and 4.2%), whereas coverage for the CWBLS method is slightly above nominal levels when using disease associated variants (between 5.9% and 10.1%).

**Table 3:** Simulation results varying the strength of the genetic predictors of the risk factor.

|  | Positive effect |  |  |  |  |  |  |  |  |
| --- | --- | --- | --- | --- | --- | --- | --- | --- | --- |
| Method | Stronger instrument |  |  | Moderate instrument |  |  | Weaker instrument |  |  |
|  | Median | SE | Power | Median | SE | Power | Median | SE | Power |
| Ratio estimate, full sample | 0.103 | 0.004 | 100.0 | 0.103 | 0.007 | 100.0 | 0.102 | 0.013 | 100.0 |
| Ratio estimate, selected sample | 0.071 | 0.010 | 100.0 | 0.072 | 0.016 | 98.6 | 0.071 | 0.031 | 65.0 |
| Inverse-probability weighting | 0.089 | 0.013 | 100.0 | 0.091 | 0.020 | 99.4 | 0.089 | 0.037 | 67.8 |
| Slope-Hunter | 0.092 | 0.018 | 75.2 | 0.085 | 0.024 | 71.5 | 0.086 | 0.037 | 65.7 |
| Multivariable IVW (IVs only) | 0.104 | 0.167 | 7.4 | 0.107 | 0.170 | 7.7 | 0.094 | 0.174 | 6.7 |
| Corrected weighted bivariate (IVs) | 0.018 | 1.729 | 3.6 | 0.036 | 1.909 | 3.2 | 0.040 | 1.806 | 2.6 |
| Multivariable IVW (including disease) | 0.105 | 0.012 | 100.0 | 0.105 | 0.018 | 100.0 | 0.105 | 0.032 | 90.0 |
| Corrected weighted bivariate (disease) | 0.105 | 0.012 | 100.0 | 0.106 | 0.017 | 100.0 | 0.106 | 0.029 | 91.2 |
|  | Null effect |  |  |  |  |  |  |  |  |
| Method | Stronger instrument |  |  | Moderate instrument |  |  | Weaker instrument |  |  |
|  | Median | SE | Coverage | Median | SE | Coverage | Median | SE | Coverage |
| Ratio estimate, full sample | 0.000 | 0.004 | 5.9 | 0.000 | 0.007 | 4.9 | 0.000 | 0.013 | 4.6 |
| Ratio estimate, selected sample | -0.029 | 0.010 | 82.6 | -0.028 | 0.016 | 41.6 | -0.029 | 0.031 | 13.9 |
| Inverse-probability weighting | -0.015 | 0.013 | 22.2 | -0.016 | 0.020 | 12.7 | -0.016 | 0.036 | 7.0 |
| Slope-Hunter | 0.004 | 0.020 | 60.8 | 0.000 | 0.024 | 52.7 | 0.001 | 0.038 | 46.2 |
| Multivariable IVW (IVs only) | -0.014 | 0.168 | 3.1 | 0.004 | 0.171 | 3.5 | -0.001 | 0.172 | 3.9 |
| Corrected weighted bivariate (IVs) | -0.037 | 1.952 | 2.0 | -0.052 | 2.055 | 2.2 | -0.053 | 1.705 | 2.4 |
| Multivariable IVW (including disease) | 0.001 | 0.012 | 3.9 | 0.000 | 0.018 | 3.7 | 0.000 | 0.032 | 3.8 |
| Corrected weighted bivariate (disease) | 0.001 | 0.012 | 7.5 | 0.000 | 0.017 | 7.5 | 0.000 | 0.029 | 7.0 |

**Table 4:** Simulation results varying the number of genetic predictors of the risk factor.

| Method | Positive effect |  |  |  |  |  |  |  |  |  |  |
| --- | --- | --- | --- | --- | --- | --- | --- | --- | --- | --- | --- |
|  | 5 instruments |  |  |  | 10 instruments |  |  |  | 20 instruments |  |  |
|  | Median | SE | Power |  | Median | SE | Power |  | Median | SE | Power |
| Ratio estimate, full sample | 0.103 | 0.009 | 100.0 |  | 0.103 | 0.007 | 100.0 |  | 0.103 | 0.005 | 100.0 |
| Ratio estimate, selected sample | 0.072 | 0.023 | 88.8 |  | 0.072 | 0.016 | 98.6 |  | 0.071 | 0.012 | 100.0 |
| Inverse-probability weighting | 0.088 | 0.028 | 90.7 |  | 0.091 | 0.020 | 99.4 |  | 0.089 | 0.015 | 100.0 |
| Slope-Hunter | 0.094 | 0.033 | 66.9 |  | 0.085 | 0.024 | 71.5 |  | 0.094 | 0.016 | 73.9 |
| Multivariable IVW (IVs only) | 0.100 | 0.274 | 4.5 |  | 0.107 | 0.170 | 7.7 |  | 0.103 | 0.115 | 13.8 |
| Corrected weighted bivariate (IVs) | 0.056 | 0.594 | 9.4 |  | 0.036 | 1.909 | 3.2 |  | -0.005 | 2.233 | 1.8 |
| Multivariable IVW (including disease) | 0.105 | 0.025 | 98.5 |  | 0.105 | 0.018 | 100.0 |  | 0.104 | 0.014 | 100.0 |
| Corrected weighted bivariate (disease) | 0.105 | 0.023 | 84.7 |  | 0.106 | 0.017 | 100.0 |  | 0.104 | 0.013 | 100.0 |
| Method | Null effect |  |  |  |  |  |  |  |  |  |  |
|  | 5 instruments |  |  |  | 10 instruments |  |  |  | 20 instruments |  |  |
|  | Median | SE | Coverage |  | Median | SE | Coverage |  | Median | SE | Coverage |
| Ratio estimate, full sample | -0.001 | 0.009 | 5.0 |  | 0.000 | 0.007 | 4.9 |  | 0.000 | 0.005 | 4.5 |
| Ratio estimate, selected sample | -0.029 | 0.023 | 23.9 |  | -0.028 | 0.016 | 41.6 |  | -0.029 | 0.012 | 70.9 |
| Inverse-probability weighting | -0.017 | 0.027 | 8.2 |  | -0.016 | 0.020 | 12.7 |  | -0.016 | 0.015 | 19.3 |
| Slope-Hunter | -0.006 | 0.035 | 50.2 |  | 0.000 | 0.024 | 52.7 |  | 0.009 | 0.016 | 63.7 |
| Multivariable IVW (IVs only) | -0.003 | 0.275 | 3.6 |  | 0.004 | 0.171 | 3.5 |  | -0.007 | 0.114 | 3.7 |
| Corrected weighted bivariate (IVs) | -0.033 | 0.629 | 7.0 |  | -0.052 | 2.055 | 2.2 |  | -0.037 | 2.097 | 1.0 |
| Multivariable IVW (including disease) | -0.001 | 0.025 | 4.0 |  | 0.000 | 0.018 | 3.7 |  | 0.000 | 0.014 | 4.0 |
| Corrected weighted bivariate (disease) | -0.001 | 0.025 | 10.1 |  | 0.000 | 0.017 | 7.5 |  | 0.000 | 0.013 | 5.9 |

The most promising methods in our simulation are inverse-probability weighting (reasonably powered, but potentially over-confident and requires individual-level data) and multivariable IVW including genetic predictors of the disease event (reasonably powered and adequate coverage under the null, but requires additional variants).

Findings from secondary scenarios are presented in the Supplementary Material:

- When varying the direction of confounding, the direction of index event bias changed, but Type 1 error inflation in the naive case-only analysis remained (Supplementary Table S1). Generally speaking, the pattern of findings was consistent across scenarios, matching the headline findings from the original setting: the inverse-probability weighting method was well-powered, although the degree of Type 1 error inflation from the method varied; the multivariable methods were well powered when including disease associated variants, but had low power when including the instrumental variants only; and the Slope-Hunter method had severely inflated Type 1 error rates in all cases.
- When varying the structure of confounding, the magnitude of index event bias varied sharply. The ratio method ignoring selection had Type 1 error rates around 100% under shared positive confounding, and close to no bias (Type 1 error rate of 6.0%) for shared negative confounding plus no additional confounding (Supplementary Table S2). However, the scenario of shared positive confounding is likely to be common in practice. Otherwise, performance of the methods was similar to the original scenario, with the inverse-probability weighting method generally reducing bias but having inflated Type 1 error rates, the Slope-Hunter method having severely inflated Type 1 error rates in all cases, and the multivariable methods giving good results when including disease associated variants, but low power otherwise. Slight Type 1 error rate inflation (6.5% to 8.9%) was observed for the CWBLS method when including disease associated variants.
- When adding a gene–confounder interaction to the effect of the genetic variants on the exposure, results were similar for the individual-level data methods, but worse for the summarized data methods as the interaction leads to bias in the summarized data (Supplementary Table S3). These methods gave biased estimates and inflated Type 1 error rates when there was an interaction term between the genetic variants and the confounder for the exposure and the time-to-event outcome, but not when there was an interaction term between the genetic variants and the confounder for the exposure and the disease event only.
- When providing the Slope-Hunter method with additional ‘genome-wide’ significant hits, the method performed better in terms of power, but still had highly inflated Type 1 error rates (over 50% in all cases, Supplementary Table S4). The multivariable IVW method performed well even when there were only 2 genetic variants associated with the disease event, whereas the CWBLS method performed badly in this case.
- When performing Heckman’s method with a binary outcome (Supplementary Table S5), the method had reasonable power with a positive effect and correct Type 1 error rate under the null. However, the method relies on the additional variants being valid instruments for the disease event. Power to detect a causal effect was similar for Heckman’s method as for the multivariable IVW method.
- When the additional variants had direct effects not only on the disease event, but also on the time-to-event outcome, the Slope-Hunter and multivariable methods gave biased estimates (Supplementary Table S6). This indicates that pleiotropy leads to bias even if the genetic predictors of the risk factor are valid instruments, and only the additional variants have direct effects on the time-to-event outcome. Similar findings were observed for Heckman’s method when the additional variants had direct effects on the outcome (results not shown).

We emphasize that this simulation study is not designed to be realistic. It informs us about the relative performance of methods, not their absolute performance, and it does this under ‘ideal’ conditions (that is, the assumptions of the methods are satisfied). For any given example, it may be that index event bias is limited in magnitude, it may be that the Type 1 error inflation due to misspecification of the inverse-probability weighting model is limited, and it may be that the power of the multivariable IVW method is low due to inadequate numbers or strength of distinct variants predicting the disease event. We only considered situations with relatively strong instruments, and so the potential benefits of the CWBLS method over the multivariable IVW method with weak instruments were not seen.

## 4 Applied examples

We proceed to explore the practical utility of these methods in two applied analyses. We consider the effects of two exposures on risk of severe COVID-19 outcomes following SARS-CoV-2 infection. High body mass index (BMI) is widely believed to increase the severity of COVID-19 outcomes, based on scientific understanding of disease pathology [50], as well as observational epidemiology [51] and Mendelian randomization investigations [52]. Interleukin-6 receptor (IL6R) inhibition is known from clinical trials to reduce the risk of severe COVID-19 outcomes [53]. We perform Mendelian randomization analyses to assess the effects of BMI and IL6R inhibition on COVID-19 progression using summarized and individual-level data.

### 4.1 Datasets for summarized data analyses

As summarized data, we use genetic associations with COVID-19 risk published by the COVID-19 Host Genetics Initiative [54]. We compare results using three GWAS datasets:

- any positive SARS-CoV-2 test versus general population (COVID-19 versus population)
- hospitalized COVID-19 versus population
- hospitalized COVID-19 versus non-hospitalized COVID-19

The last of these datasets should provide estimates that are subject to index event bias, as the controls for this dataset were selected as those with a positive SARS-CoV-2 test. We think of hospitalized COVID-19 as representing disease progression, as cases have had an initial disease infection event, which then progressed to a more serious disease state.

For BMI, we take genetic associations published by Pulit et al. [55]. Estimates are presented as odds ratios (OR) per 1 standard deviation (approximately 4.8 kg/m^2^) increase in genetically-predicted BMI. For IL6R inhibition, C-reactive protein (CRP) is used as a biomarker of this mechanism. We use CRP as the exposure in our analyses for IL6R inhibition, as has been done previously [56], and take genetic associations with CRP published by Said et al. [57]. Estimates are presented as OR per unit increase in log-transformed genetically-predicted CRP levels.

### 4.2 Dataset for individual-level data analyses

All individual-level data analyses were conducted in UK Biobank, a longitudinal cohort study of around 500,000 people from the UK aged 40 to 69 years at baseline, recruited between 2006 and 2010 [58]. We restricted analyses to individuals of European ancestry as determined based on UK Biobank genotype data using fine-scale ancestry inference [59], retaining participants with *>* 90% European ancestry defined as the sum of ancestry components AC23 (UK/Ireland) and AC34 (other European) for each participant. We additionally excluded participants who did not achieve whole-genome sequencing quality requirements (UK Biobank Data-Field 32064), and those missing required genotype, outcome, or covariate data (complete-case analysis for age, sex, BMI, systolic blood pressure, Townsend deprivation index, smoking status, C-reactive protein, and genetic principal components).

The incident disease outcome was defined as having a recorded electronic health record event with COVID-19 as the main International Statistical Classification of Diseases 10th Revision (ICD-10) diagnosis (ICD-10 U07.*). The progression outcome was defined as death with COVID-19 listed as the underlying (primary) cause of death (ICD-10 U07.*).

After applying all restrictions, the BMI analysis included 418 865 participants, with 4 269 incident COVID-19 events and 1 234 COVID-19 deaths. In the IL6R analysis (using the corresponding IL6R instrument and the same outcome definitions and exclusions), the final dataset included 398 262 participants, with 4 043 incident COVID-19 events and 1 179 COVID-19 deaths.

### 4.3 Genetic variants

For BMI, we take all variants associated with BMI in Pulit et al. at a GWAS significance level (*p <* 5 *×* 10^−8^) after harmonization and variant-level quality control as instrumental variants (542 variants). For IL6R inhibition, we take rs2228145 as the sole instrumental variant; this is a missense variant in the *IL6R* gene region that has previously been used as an instrument for IL6R inhibition, and associates with circulating CRP levels [60]. In the multivariable methods for IL6R, we additionally include the top five variants associated with COVID-19 incidence at a GWAS significance level (*p <* 5 *×* 10^−8^) after pruning at *r*^2^ *<* 0.01 taken from the COVID-19 versus population dataset.

### 4.4 Statistical analyses

We compare six analysis strategies with summarized data:

- *Effect on incidence:* Univariable IVW (multiple variants) / ratio method (single variant) with outcome associations from the COVID-19 versus population dataset – this analysis relates to the effect of the risk factor on incident disease risk.
- *Effect on progression (no index event bias):* Univariable IVW (multiple variants) / ratio method (single variant) with outcome associations from the hospitalized COVID-19 versus population dataset – this analysis should not be subject to index event bias, as controls are taken from the general population.
- *Effect on progression (index event bias):* Univariable IVW (multiple variants) / ratio method (single variant) with outcome associations from the hospitalized COVID-19 versus non-hospitalized COVID-19 dataset – this analysis should be subject to index event bias, as controls are selected as those with a positive COVID-19 test.
- Slope-Hunter with outcome associations from the hospitalized COVID-19 versus non-hospitalized COVID-19 dataset and disease event associations from the COVID-19 versus population dataset.
- Multivariable IVW with outcome associations from the hospitalized COVID-19 versus non-hospitalized COVID-19 dataset and adjustment for associations from the COVID-19 versus population dataset.
- CWBLS with outcome associations from the hospitalized COVID-19 versus non-hospitalized COVID-19 dataset and adjustment for associations from the COVID-19 versus population dataset.

We also perform two analyses with individual-participant data. We initially regress the risk factor on the instrumental variants to obtain genetically-predicted values of the risk factor, and then regress the outcome on genetically-predicted values of the risk factor:

- *Two-stage method:* We regress the progression outcome (that is, COVID-19 death) on genetically-predicted values of the risk factor with adjustment for age, sex, and 10 genomic principal components in a logistic regression model in participants with the incident event (that is, any recorded COVID-19 event).
- *Two-stage with inverse-probability weighting:* We perform the same logistic regression in participants with the incident event, but weighting participants based on the inverse of their propensity score for the incident event. The propensity score model was a logistic regression of the incident event on the risk factor, genetic instruments, age, age-squared, sex, BMI, systolic blood pressure, and Townsend deprivation index.

We note that the summarized and individual-level data analyses are based on different datasets and have different definitions of the progression outcome, and so caution is required when comparing estimates between the approaches.

### 4.5 Results

Results are shown in Figure 3 (BMI) and Figure 4 (IL6R). For BMI, all estimates are positive, and most are statistically significant at the conventional *p <* 0.05 threshold. There is evidence for an effect of BMI on COVID-19 incidence (OR 1.17, 95% confidence interval [CI] 1.14, 1.20), and on COVID-19 progression from the dataset that is not subject to index event bias of hospitalized COVID-19 versus population (OR 1.51, 95% CI 1.43, 1.60). In the dataset subject to index event bias (hospitalized COVID-19 versus non-hospitalized COVID-19), the estimate is attenuated towards the null, but we still see evidence for an effect of BMI on COVID-19 progression (OR 1.35, 95% CI 1.21, 1.50). This suggests that, while this analysis is subject to bias, the bias does not materially affect findings in terms of their direction for this example. Methods that attempt to account for index event bias did not correct estimates in the direction of the gold-standard estimate.

**Figure 3.**
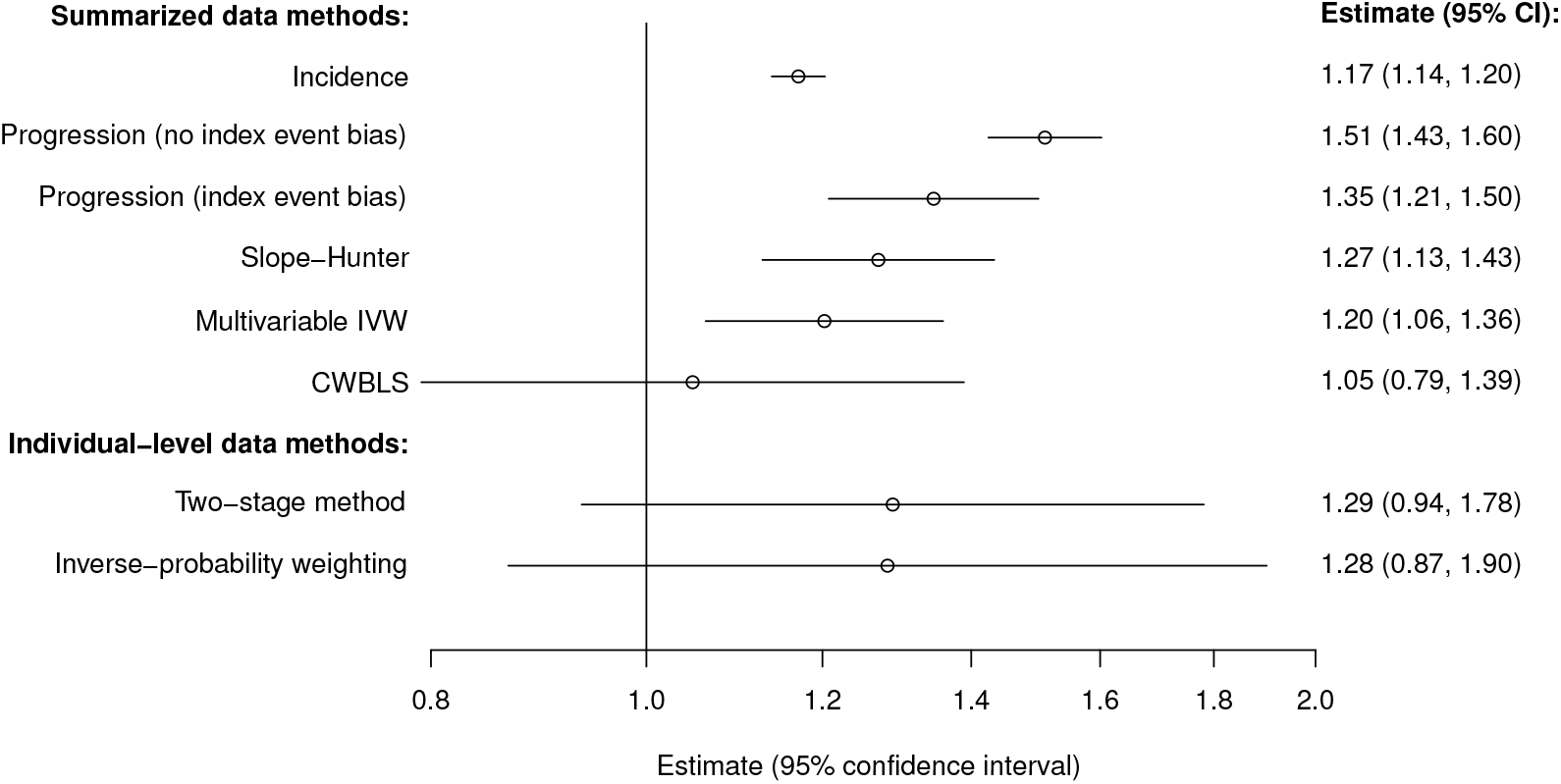
Mendelian randomization estimates for body mass index (BMI) on COVID-19 risk and progression. Estimates represent odds ratios per 1 standard deviation higher genetically-predicted BMI. Note that the summarized and individual-level analyses are based on different data sources, and have different definitions of the progression outcome.

**Figure 4.**
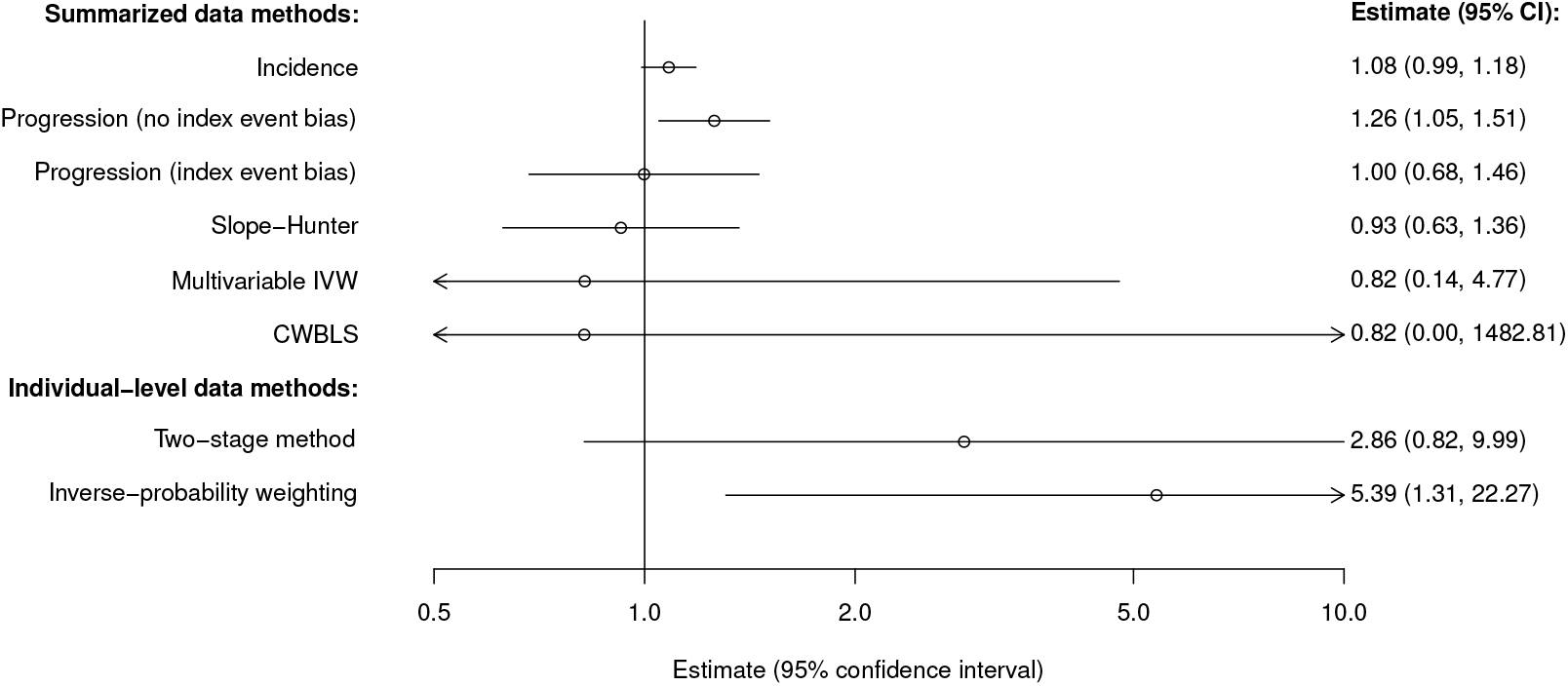
Mendelian randomization estimates for interleukin 6 receptor (IL6R) on COVID-19 risk and progression. Estimates represent odds ratios per 1 unit higher genetically-predicted log-transformed C-reactive protein. Note that the summarized and individual-level analyses are based on different data sources, and have different definitions of the progression outcome.

For IL6R inhibition, there is weak evidence for an effect on COVID-19 incidence (OR 1.08, 95% CI 0.99, 1.18), and stronger evidence for an effect on COVID-19 progression from the gold-standard dataset (OR 1.26, 95% CO 1.05, 1.51). However, in the dataset subject to index event bias, there is no clear evidence for an effect on COVID-19 progression, with the point estimate being very close to the null (OR 1.00, 95% CI 0.68, 1.46). Again, the summarized data methods that attempt to account for index event bias did not correct estimates in the direction of the gold-standard estimate. In the individual-level data analysis, the estimate without correction for index event bias was positive but non-significant (OR 2.86, 95% CI 0.82, 9.99), while the estimate with inverse-probability weighting to correct for index event bias was larger and statistically significant (OR 5.39, 95% CI 1.31, 22.27). One reason why results from the multivariable methods are disappointing is that the additional variants that affect COVID-19 incidence likely also affect COVID-19 progression directly (that is, they are pleiotropic).

## 5 Discussion

There are many factors that can bias Mendelian randomization analyses. Index event bias is perhaps less critical than bias due to pleiotropy or population stratification, but it can lead to false positive and false negative findings. For example, an early pre-print draft of work investigating potential effects of 25-hydroxyvitamin D levels (a biomarker of vitamin D levels) on risk of COVID-19 showed a positive association between genetically-predicted 25-hydroxyvitamin D and COVID-19 risk, suggesting that vitamin D supplementation may increase risk of COVID-19 [61]. However, this analysis was based on an outcome dataset that is subject to index event bias. Conversely, there will also be cases where there is index event bias, but it does not change findings in a material way, such as the analyses for BMI on COVID-19 risk presented here.

While statistical methods have been developed to combat index event bias, they all have some deficiencies. We were unable to obtain sensible results from the Slope-Hunter method in any simulation scenario, and in applied analyses, it did not reduce index event bias in any analysis. Heckman’s method is not particularly flexible, and depends on the availability of instruments for the disease event. For multivariable methods, including the standard multivariable IVW method and the CWBLS method, we obtained reasonable results in simulations when we had variants that affect incident disease risk in a specific way. However, in the absence of such variants, estimates were imprecise, and if variants that affect disease risk also affect disease progression, bias could be severe. For the inverse-probability weighting method, in order to obtain reasonable results, we require individual-level data and a statistical model that accurately predicts incident disease risk.

For many diseases, investigating causal effects of risk factors on disease progression is unnecessary, because the same factors that affect disease progression also affect disease risk. Analyses for disease risk are not affected by index event bias, are simpler to perform, and typically have larger sample sizes. Cases where the same mechanisms (and hence the same genetic variants) affect disease risk and disease progression are not likely to be amenable to multivariable methods to correct for index event bias. However, in such cases, analyses to investigate effects on disease progression are less critical. Multivariable methods are likely to be valuable for outcomes that occur subsequent to the disease but are affected by different mechanisms, such as kidney disease in Type 1 diabetes patients. As different mechanisms are likely to affect Type 1 diabetes risk and kidney dysfunction, we are more likely to be able to find genetic instruments for disease risk that do not have direct (pleiotropic) effects on disease progression.

In other cases, inverse-probability weighting may be a worthwhile analysis strategy. If analysts do not have access to individual-level data or plausible instruments for incident disease risk, an alternative strategy is to perform a naive analysis ignoring index event bias, and to investigate the potential magnitude of index event bias in a simulation analysis. For instance, in a Mendelian randomization investigation into the effect of BMI on breast cancer progression, the association of genetically-predicted BMI with breast cancer progression was so strong that it could not plausibly be fully explained by realistic levels of index event bias [62].

There are several limitations of this investigation. All scenarios are different, and it is not possible to perform a simulation study that covers all possible cases. Additionally, we did not cover all possible methods. In some cases, robust methods for multivariable Mendelian randomization (such as the multivariable MR-Lasso and MR-Horse methods) may be worthwhile, as these methods are robust to some genetic variants having pleiotropic effects [63]. A further approach that we did not consider here when the outcome evolves over time (but stays in the same format – such as a mental health score that is measured at different timepoints) is a multivariate analysis [64].

In summary, while there are several methods to combat index event bias in Mendelian randomization, none of these are perfect solutions. A suggested analysis strategy is: 1) if the same mechanisms affect disease progression as affect disease risk, perform an analysis for disease risk rather than for disease progression; 2) if there are distinct variants affecting disease risk, consider performing a multivariable analysis with the risk factor and disease risk as exposures; 3) if you have access to individual-level data, consider performing an inverse-probability weighted analysis; and 4) if none of these approaches are possible, perform a naive analysis and estimate the likely magnitude of bias that could occur under realistically severe index event bias.

## Author contributions

Linxuan Zhang, Ixavier Alonzo Higgins, and Stephen Burgess wrote the initial draft. Stephen Burgess performed simulations. Linxuan Zhang performed applied analyses. All authors contributed to the design and implementation of the research, to the interpretation of the results, and to the drafting of the manuscript. All authors approve of the final manuscript, and the decision to submit for publication.

## Acknowledgements

This research has been conducted using the UK Biobank Resource under Application Number 536820.

## Data and code availability

Example code is included in the Supplementary Material. The complete simulation code is available at https://github.com/sb452/mrprog. UK Biobank data is available on application to any *bona fide* researcher.

## S.1 Additional simulation study scenarios

We describe the five additional simulation scenarios in detail here:

### Direction of confounding

We generate the disease event *D* as a binomial random variable:

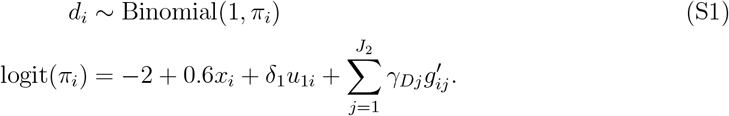

and the time-to-event outcome *Y* as an exponential random variable:

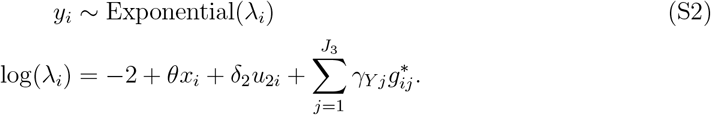

This is the same as the main simulation scenario, except that we originally set *δ*_1_ = −1 and *δ*_2_ = +1. We consider all positive signs of *δ*_1_ and *δ*_2_:

- Positive / positive (+/+): *δ*_1_ = +1 and *δ*_2_ = +1,
- Negative / positive (−/+): *δ*_1_ = −1 and *δ*_2_ = +1 (this is the original parameter setting),
- Positive / negative (+/−): *δ*_1_ = +1 and *δ*_2_ = −1,
- Negative / negative (−/−): *δ*_1_ = −1 and *δ*_2_ = −1.

### Structure of confounding

In addition to *U*_1_ and *U*_2_ (which both affect the risk factor), we generate a third confounder *U*_3_ with an independent standard normal distribution. We generate the disease event *D* as a binomial random variable:

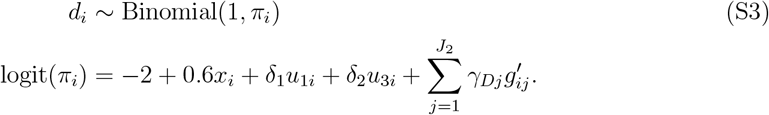

and the time-to-event outcome *Y* as an exponential random variable:

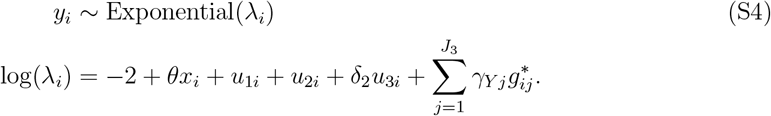

This is similar to the main simulation scenario, except that in the main scenario, *U*_1_ was a confounder of the risk factor and disease event. Here, *U*_1_ is a confounder of the risk factor, disease event, and time-to-event outcome, and *U*_3_ is a confounder of the disease event and time-to-event outcome (when *δ*_2_ ≠ 0). *U*_2_ remains as a confounder of the risk factor and time-to-event outcome in both cases. We consider four values for *δ*_1_ and *δ*_2_:

- Shared positive: *δ*_1_ = +1 and *δ*_2_ = 0,
- Shared negative: *δ*_1_ = −1 and *δ*_2_ = 0,
- Shared positive plus: *δ*_1_ = +1 and *δ*_2_ = +1,
- Shared negative plus: *δ*_1_ = −1 and *δ*_2_ = +1.

### Gene–confounder interaction

We define *g*_*sum*_ as the sum of the instrumental variants, such that *g*_*sum,i*_ = ∑_*j*_ *g*_*ij*_. We generate the exposure *X* as:

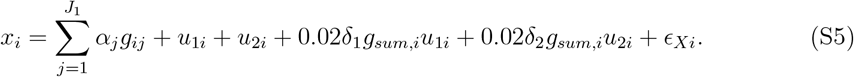

We consider three values for *δ*_1_ and *δ*_2_:

- Gene–confounder interaction for *U*_2_ only: *δ*_1_ = 0 and *δ*_2_ = +1,
- Gene–confounder interaction for *U*_1_ only: *δ*_1_ = +1 and *δ*_2_ = 0,
- Gene–confounder interaction for *U*_1_ and *U*_2_: *δ*_1_ = +1 and *δ*_2_ = +1.

### Additional ‘genome-wide’ variants

The Slope-Hunter method requires genome-wide association data to estimate the degree of collider bias. In the original simulation, we simulated data on associations for 1000 additional variants. Associations were obtained by simulating independent beta-coefficients from a normal distribution with mean 0 and standard deviation 0.2, and their standard errors from a uniform distribution between 0.15 and 0.25.

These null associations, together with the associations for the original variants, formed the inputs for the Slope-Hunter method in the main simulation scenarios.

In an additional scenario, we varied the number of truly associated variants in the additional variants, simulating 2, 5, 10 (original), and 20 variants with a direct effect on the disease event, and the same number of variants with a direct effect on the time-to-event outcome.

### Heckman’s method with additional variants

Heckman’s method can be implemented with either a normally-distributed continuous outcome or a binary outcome. We generated a binary outcome *Y* as:

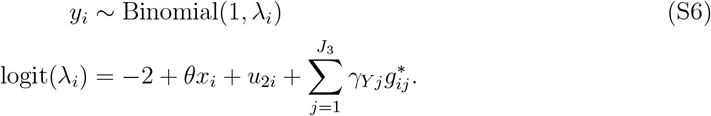

This is the same form of the linear predictor as before, except that the outcome is generated using a binomial distribution rather than an exponential distribution. We note that to force the statistical package to recognize the outcome as binary, it was necessary to recode the outcome as a logical variable (`TRUE/FALSE`). We varied the number of additional variants affecting the disease event, simulating 2, 5, 10, and 20 variants with a direct effect on the disease event. For comparison, we also perform the multivariable IVW method in this scenario, using summarized data for the instrumental variants and the additional variants. In this simulation, genetic associations with the outcome are estimated using logistic regression. We note that the quantities estimated by the two methods differ, but they should be comparable in terms of power and coverage.

### Additional variants affect the time-to-event outcome

In a further additional scenario, we simulated a single set of variants that affect both the disease event and the time-to-event outcome, by combining the 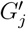 and 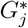 variants into a single set. We considered settings with 2, 5, 10, and 20 of these variants. When included in a multivariable model, these variants would be considered as pleiotropic, as they have direct effects on the outcome that do not operate via either of the exposures (that is, via the risk factor or the disease event).

## S.2 Illustrative simulation study code

~~~
expit <-function(x) { return(exp(x)/(1+exp(x))) }
logit <-function(x) { return(log(x/(1-x))) }
library(survival)
library(MendelianRandomization)
library(ColliderBias)
library(SlopeHunter)
library(sampleSelection)
set.seed(496)
parts = 1e5 vars = 10
vars_ex = 10
vars_slope = 1000
times = 100
alpha0 = 0.15 # moderate instrument setting
## simulation of individual-level data
g = array(rnorm(parts*(vars+2*vars_ex)), dim=c(parts, vars+2*vars_ex))
alpha = c(rnorm(vars, alpha0, 0.02), rep(0, 2*vars_ex))
gamma_y = c(rep(0, vars), rnorm(vars_ex, 0.2, 0.05), rep(0, vars_ex))
gamma_s = c(rep(0, vars), rep(0, vars_ex), rnorm(vars_ex, 0.2, 0.05))
u1 = rnorm(parts)
u2 = rnorm(parts)
x = rnorm(parts, g%*%alpha+u1+u2)
y = rbinom(parts, 1, expit(−2+0.6*x-u1+g%*%gamma_y))
gscore = g%*%c(rep(1, vars), rep(0, vars_ex*2))
s = rexp(parts, exp(−2+0.2*x+u2+g%*%gamma_s))
s1 = ifelse(y==1, s, NA)
s2 = rep(1, parts)
## calculation of summarized data
beta_s1 = NULL; se_s1 = NULL
beta_x = NULL; se_x = NULL
beta_y = NULL; se_y = NULL
for (k in 1:(vars+2*vars_ex)) {
 reg_s1 = summary(coxph(Surv(s1, s2)∼g[,k]))
 beta_s1[k] = reg_s1$coef[1,1]
 se_s1[k] = reg_s1$coef[1,3]
 reg_y = summary(glm(y∼g[,k], family=“binomial”))
 beta_y[k] = reg_y$coef[2,1]
 se_y[k] = reg_y$coef[2,2]
 reg_x = summary(lm(x∼g[,k]))
 beta_x[k] = reg_x$coef[2,1]
 se_x[k] = reg_x$coef[2,2]
 }
## naive method
cox_reg = summary(coxph(Surv(s, s2)∼gscore)) # all individuals
cox1_reg = summary(coxph(Surv(s1, s2)∼gscore)) # diseased individuals only
## bivariate method
mv1_reg = mr_mvivw(mr_mvinput(cbind(beta_y[1:vars],
 beta_x[1:vars]),bind(se_y[1:vars], se_x[1:vars]),
 beta_s1[1:vars], se_s1[1:vars]))
# instrumental variants only
mvx1_reg = mr_mvivw(mr_mvinput(cbind(beta_y[1:(vars+vars_ex)],
 beta_x[1:(vars+vars_ex)]), cbind(se_y[1:(vars+vars_ex)], se_x[1:(vars+vars_ex)]),
 beta_s1[1:(vars+vars_ex)], se_s1[1:(vars+vars_ex)]))
# including disease event variants
## IPW method
prop_score = glm(y∼x+gscore+x*gscore, family=“binomial”)$fitted
ipw1_reg = summary(coxph(Surv(s1, s2)∼gscore, weights = prop_score^-1))
## Heckman’s method
heck1_reg = summary(selection(y∼g[,(vars+1):(vars+vars_ex)]+gscore, s1∼gscore))
# this only works for a normally-distributed continuous outcome or a binary outcome
# for a binary outcome, recode s1 as TRUE / FALSE
## corrected weighted least squares
summ.data1 = data.frame(xbeta=beta_x[1:vars], xse=se_x[1:vars], dbeta=beta_y[1:vars], dse=se_y[1:vars], ybeta=beta_s1[1:vars], yse=se_s1[1:vars])
CWBLS1 = CWBLS(summ.data1)
# instrumental variants only
summ.data1x = data.frame(xbeta=beta_x[1:(vars+vars_ex)], xse=se_x[1:(vars+vars_ex)],
 dbeta=beta_y[1:(vars+vars_ex)], dse=se_y[1:(vars+vars_ex)],
 ybeta=beta_s1[1:(vars+vars_ex)], yse=se_s1[1:(vars+vars_ex)])
CWBLS1x = CWBLS(summ.data1x)
# including disease event variants
## slope hunter
# generating simulated GWAS data
betax_s1 = rnorm(vars_slope, 0, 0.2); sex_s1 = runif(vars_slope, 0.15, 0.25)
betax_s0 = rnorm(vars_slope, 0, 0.2); sex_s0 = runif(vars_slope, 0.15, 0.25)
betax_y = rnorm(vars_slope, 0, 0.2); sex_y = runif(vars_slope, 0.15, 0.25)
beta_s1plus = c(beta_s1, betax_s1); se_s1plus = c(se_s1, sex_s1)
beta_yplus = c(beta_y, betax_y); se_yplus = c(se_y, sex_y)
dat_assocs = data.frame(beta_s1plus, se_s1plus, beta_yplus, se_yplus)
hunt1 = hunt(dat=dat_assocs, xbeta_col = “beta_yplus”, xse_col = “se_yplus”,
 ybeta_col = “beta_s1plus”, yse_col = “se_s1plus”, show_adjustments=TRUE, Plot=FALSE, M=100)
beta_s1hunt = hunt1$est$ybeta_adj[1:vars]
 se_s1hunt = hunt1$est$yse_adj[1:vars]
ivw_hunt1 = mr_ivw(mr_input(beta_x[1:vars], se_x[1:vars], beta_s1hunt, se_s1hunt))
~~~

**Supplementary Table S1:**
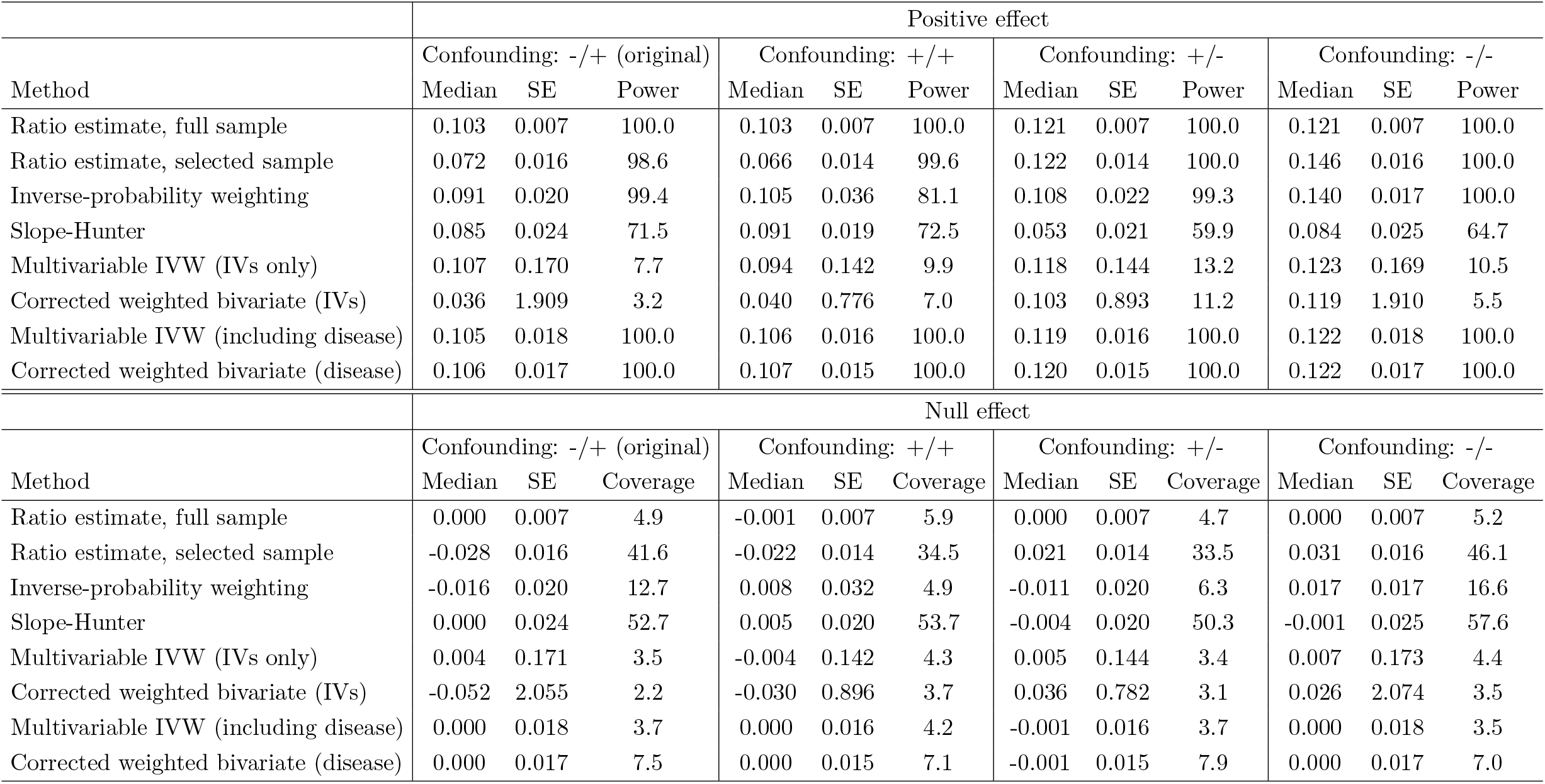
Additional simulation results varying the direction of confounding. The leftmost columns (confounding: −/+) represents the original simulation setting.

**Supplementary Table S2:**
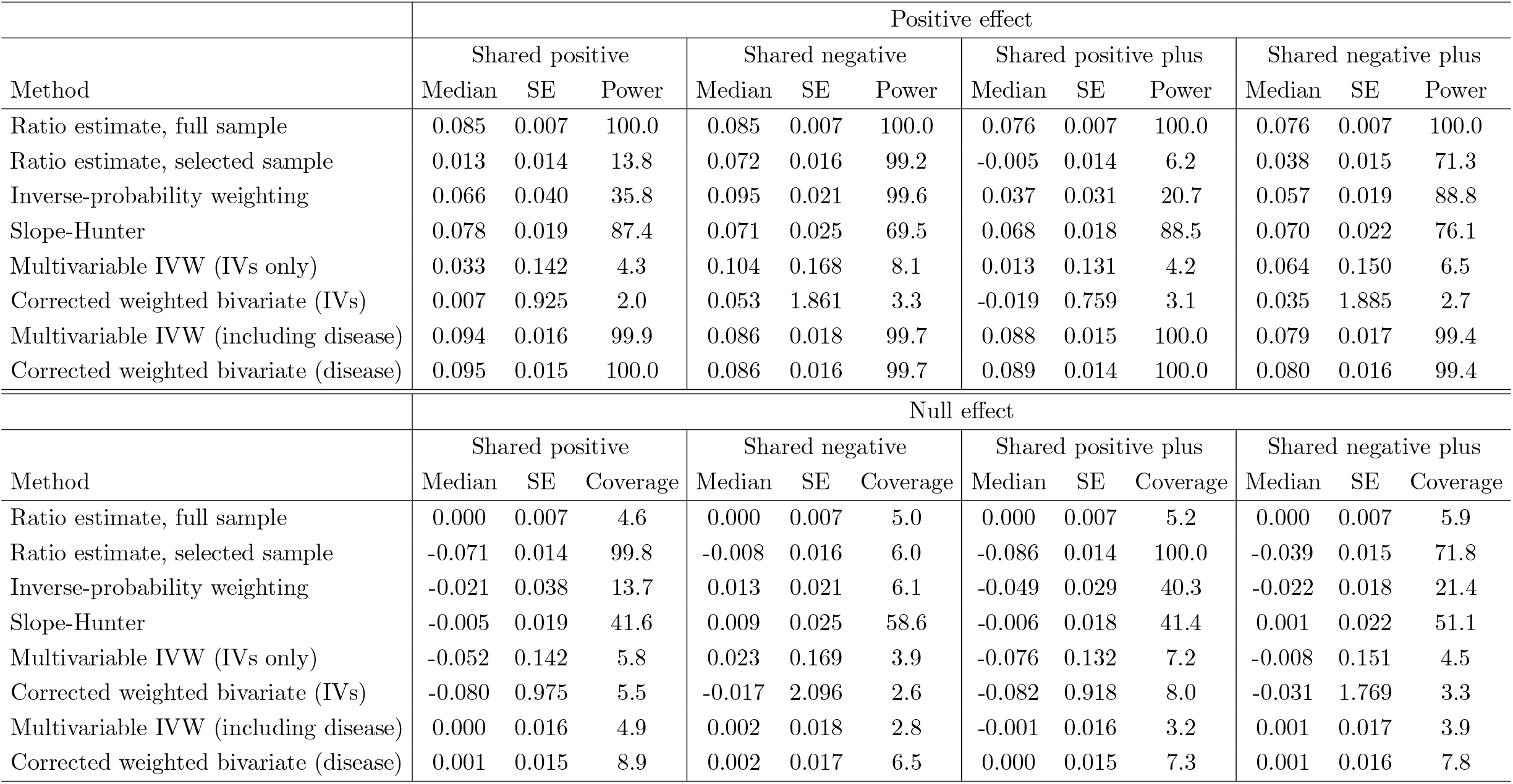
Additional simulation results varying the structure of confounding.

| Method | Positive effect |  |  |  |  |  |  |  |  |  |  |  |
| --- | --- | --- | --- | --- | --- | --- | --- | --- | --- | --- | --- | --- |
|  | Shared positive |  |  | Shared negative |  |  | Shared positive plus |  |  | Shared negative plus |  |  |
|  | Median | SE | Power | Median | SE | Power | Median | SE | Power | Median | SE | Power |
| Ratio estimate, full sample | 0.085 | 0.007 | 100.0 | 0.085 | 0.007 | 100.0 | 0.076 | 0.007 | 100.0 | 0.076 | 0.007 | 100.0 |
| Ratio estimate, selected sample | 0.013 | 0.014 | 13.8 | 0.072 | 0.016 | 99.2 | -0.005 | 0.014 | 6.2 | 0.038 | 0.015 | 71.3 |
| Inverse-probability weighting | 0.066 | 0.040 | 35.8 | 0.095 | 0.021 | 99.6 | 0.037 | 0.031 | 20.7 | 0.057 | 0.019 | 88.8 |
| Slope-Hunter | 0.078 | 0.019 | 87.4 | 0.071 | 0.025 | 69.5 | 0.068 | 0.018 | 88.5 | 0.070 | 0.022 | 76.1 |
| Multivariable IVW (IVs only) | 0.033 | 0.142 | 4.3 | 0.104 | 0.168 | 8.1 | 0.013 | 0.131 | 4.2 | 0.064 | 0.150 | 6.5 |
| Corrected weighted bivariate (IVs) | 0.007 | 0.925 | 2.0 | 0.053 | 1.861 | 3.3 | -0.019 | 0.759 | 3.1 | 0.035 | 1.885 | 2.7 |
| Multivariable IVW (including disease) | 0.094 | 0.016 | 99.9 | 0.086 | 0.018 | 99.7 | 0.088 | 0.015 | 100.0 | 0.079 | 0.017 | 99.4 |
| Corrected weighted bivariate (disease) | 0.095 | 0.015 | 100.0 | 0.086 | 0.016 | 99.7 | 0.089 | 0.014 | 100.0 | 0.080 | 0.016 | 99.4 |
| Method | Null effect |  |  |  |  |  |  |  |  |  |  |  |
|  | Shared positive |  |  | Shared negative |  |  | Shared positive plus |  |  | Shared negative plus |  |  |
|  | Median | SE | Coverage | Median | SE | Coverage | Median | SE | Coverage | Median | SE | Coverage |
| Ratio estimate, full sample | 0.000 | 0.007 | 4.6 | 0.000 | 0.007 | 5.0 | 0.000 | 0.007 | 5.2 | 0.000 | 0.007 | 5.9 |
| Ratio estimate, selected sample | -0.071 | 0.014 | 99.8 | -0.008 | 0.016 | 6.0 | -0.086 | 0.014 | 100.0 | -0.039 | 0.015 | 71.8 |
| Inverse-probability weighting | -0.021 | 0.038 | 13.7 | 0.013 | 0.021 | 6.1 | -0.049 | 0.029 | 40.3 | -0.022 | 0.018 | 21.4 |
| Slope-Hunter | -0.005 | 0.019 | 41.6 | 0.009 | 0.025 | 58.6 | -0.006 | 0.018 | 41.4 | 0.001 | 0.022 | 51.1 |
| Multivariable IVW (IVs only) | -0.052 | 0.142 | 5.8 | 0.023 | 0.169 | 3.9 | -0.076 | 0.132 | 7.2 | -0.008 | 0.151 | 4.5 |
| Corrected weighted bivariate (IVs) | -0.080 | 0.975 | 5.5 | -0.017 | 2.096 | 2.6 | -0.082 | 0.918 | 8.0 | -0.031 | 1.769 | 3.3 |
| Multivariable IVW (including disease) | 0.000 | 0.016 | 4.9 | 0.002 | 0.018 | 2.8 | -0.001 | 0.016 | 3.2 | 0.001 | 0.017 | 3.9 |
| Corrected weighted bivariate (disease) | 0.001 | 0.015 | 8.9 | 0.002 | 0.017 | 6.5 | 0.000 | 0.015 | 7.3 | 0.001 | 0.016 | 7.8 |

**Supplementary Table S3:**
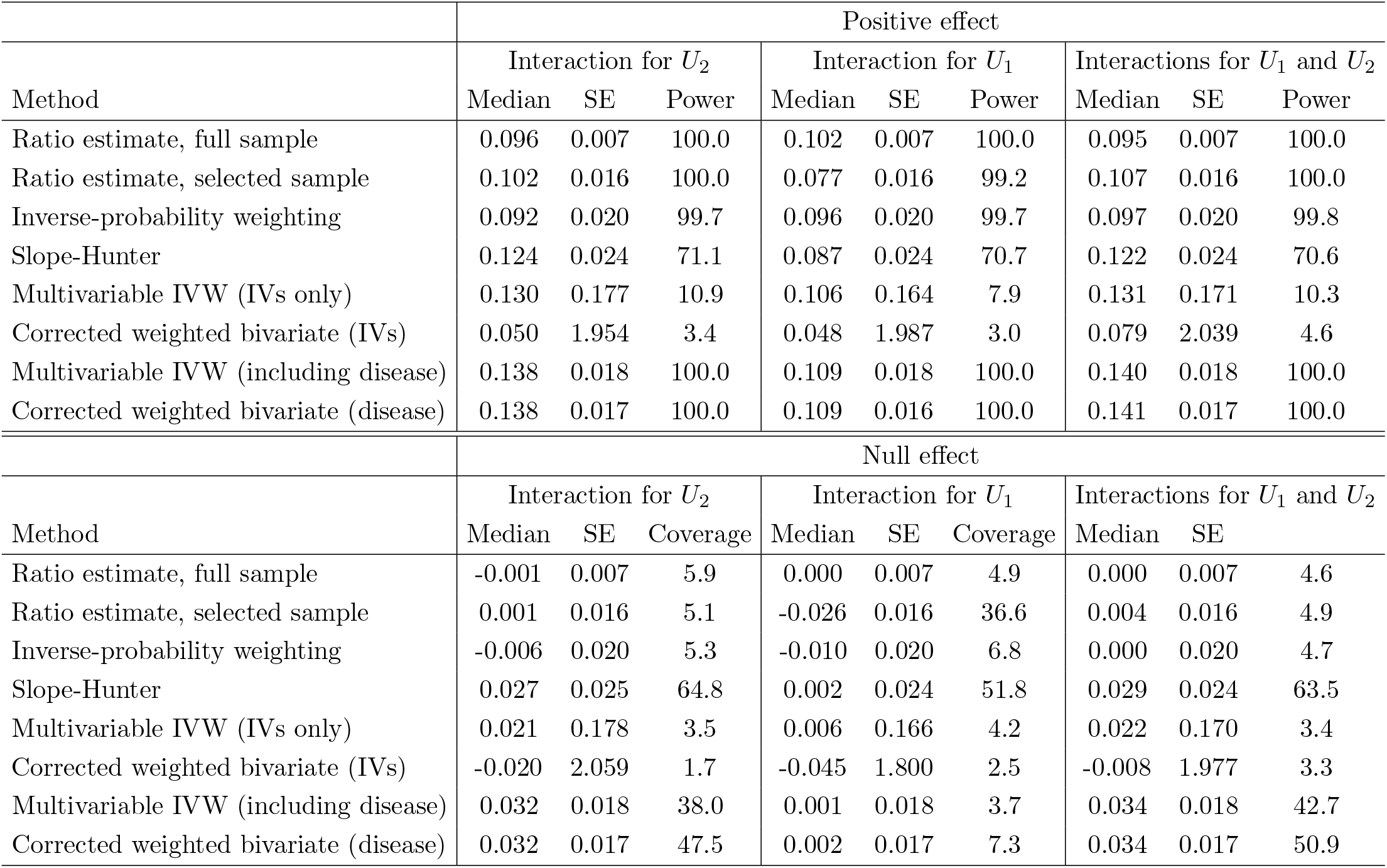
Additional simulation results adding gene–confounder interaction.

**Supplementary Table S4:** Simulation results varying the number of additional ‘genome-wide’ hits provided to the Slope-Hunter and multivariable methods (multivariable inverse variance weighted, MVIVW; and corrected weighted bivariate least squares, CWBLS).

| Method | Positive effect |  |  |  |  |  |  |  |  |  |  |  |
| --- | --- | --- | --- | --- | --- | --- | --- | --- | --- | --- | --- | --- |
|  | 2 additional hits |  |  | 5 additional hits |  |  | 10 additional hits (original) |  |  | 20 additional hits |  |  |
|  | Median | SE | Power | Median | SE | Power | Median | SE | Power | Median | SE | Power |
| Ratio estimate, full sample | 0.109 | 0.007 | 100.0 | 0.106 | 0.007 | 100.0 | 0.103 | 0.007 | 100.0 | 0.096 | 0.007 | 100.0 |
| Slope-Hunter | 0.016 | 0.023 | 65.5 | 0.084 | 0.023 | 70.9 | 0.085 | 0.024 | 71.5 | 0.089 | 0.022 | 74.4 |
| MVIVW (including disease) | 0.110 | 0.025 | 98.5 | 0.109 | 0.020 | 100.0 | 0.105 | 0.018 | 100.0 | 0.097 | 0.017 | 100.0 |
| CWBLS (disease) | 0.111 | 0.234 | 14.2 | 0.109 | 0.019 | 93.7 | 0.106 | 0.017 | 100.0 | 0.097 | 0.016 | 100.0 |
| Method | Null effect |  |  |  |  |  |  |  |  |  |  |  |
|  | 2 additional hits |  |  | 5 additional hits |  |  | 10 additional hits (original) |  |  | 20 additional hits |  |  |
|  | Median | SE | Coverage | Median | SE | Coverage | Median | SE | Coverage | Median | SE | Coverage |
| Ratio estimate, full sample | 0.000 | 0.007 | 5.0 | 0.000 | 0.007 | 5.2 | 0.000 | 0.007 | 4.9 | 0.000 | 0.007 | 4.6 |
| Slope-Hunter | 0.005 | 0.023 | 61.2 | 0.008 | 0.024 | 58.8 | 0.000 | 0.024 | 52.7 | -0.001 | 0.023 | 58.7 |
| MVIVW (including disease) | -0.001 | 0.025 | 3.9 | 0.000 | 0.020 | 5.2 | 0.000 | 0.018 | 3.7 | 0.000 | 0.017 | 4.0 |
| CWBLS (disease) | -0.001 | 0.223 | 0.1 | 0.000 | 0.020 | 7.4 | 0.000 | 0.017 | 7.5 | 0.000 | 0.016 | 8.6 |

**Supplementary Table S5:** Simulation results with a binary outcome varying the number of instruments for the disease event provided to Heckman’s method and the multivariable inverse variance weighted (MVIVW) method.

|  | Positive effect |  |  |  |  |  |  |  |  |  |  |  |
| --- | --- | --- | --- | --- | --- | --- | --- | --- | --- | --- | --- | --- |
|  | 2 additional hits |  |  | 5 additional hits |  |  | 10 additional hits |  |  | 20 additional hits |  |  |
| Method | Median | SE | Power | Median | SE | Power | Median | SE | Power | Median | SE | Power |
| Heckman's | 0.014 | 0.005 | 81.6 | 0.014 | 0.004 | 93.2 | 0.013 | 0.004 | 95.2 | 0.013 | 0.003 | 96.8 |
| MVIVW (including disease) | 0.159 | 0.058 | 75.7 | 0.157 | 0.047 | 91.2 | 0.152 | 0.042 | 94.0 | 0.148 | 0.038 | 96.5 |
|  | Null effect |  |  |  |  |  |  |  |  |  |  |  |
|  | 2 additional hits |  |  | 5 additional hits |  |  | 10 additional hits |  |  | 20 additional hits |  |  |
| Method | Median | SE | Coverage | Median | SE | Coverage | Median | SE | Coverage | Median | SE | Coverage |
| Heckman's | 0.000 | 0.005 | 5.1 | 0.000 | 0.004 | 4.8 | 0.000 | 0.004 | 6.8 | 0.000 | 0.003 | 4.7 |
| MVIVW (including disease) | -0.001 | 0.061 | 3.9 | 0.001 | 0.049 | 3.8 | 0.001 | 0.044 | 6.3 | -0.002 | 0.040 | 4.1 |

**Supplementary Table S6:** Simulation results when the additional ‘genome-wide’ hits provided to the Slope-Hunter and multivariable methods (multivariable inverse variance weighted, MVIVW; and corrected weighted bivariate least squares, CWBLS) have direct effects on the time-to-event outcome.

| Method | Positive effect |  |  |  |  |  |  |  |  |  |  |  |
| --- | --- | --- | --- | --- | --- | --- | --- | --- | --- | --- | --- | --- |
|  | 2 additional hits |  |  | 5 additional hits |  |  | 10 additional hits |  |  | 20 additional hits |  |  |
|  | Median | SE | Power | Median | SE | Power | Median | SE | Power | Median | SE | Power |
| Ratio estimate, full sample | 0.109 | 0.007 | 100.0 | 0.107 | 0.007 | 100.0 | 0.103 | 0.007 | 100.0 | 0.096 | 0.007 | 100.0 |
| Slope-Hunter | -0.010 | 0.028 | 53.7 | -0.012 | 0.029 | 62.9 | -0.039 | 0.032 | 73.1 | -0.155 | 0.028 | 88.3 |
| MVIVW (including disease) | -0.215 | 0.039 | 95.1 | -0.199 | 0.054 | 93.9 | -0.193 | 0.063 | 93.8 | -0.185 | 0.066 | 94.9 |
| CWBLS (disease) | -0.221 | 0.256 | 16.0 | -0.202 | 0.056 | 87.8 | -0.195 | 0.039 | 100.0 | -0.187 | 0.029 | 99.9 |
| Method | Null effect |  |  |  |  |  |  |  |  |  |  |  |
|  | 2 additional hits |  |  | 5 additional hits |  |  | 10 additional hits |  |  | 20 additional hits |  |  |
|  | Median | SE | Coverage | Median | SE | Coverage | Median | SE | Coverage | Median | SE | Coverage |
| Ratio estimate, full sample | 0.000 | 0.007 | 4.3 | 0.000 | 0.007 | 5.4 | 0.000 | 0.007 | 3.8 | 0.000 | 0.007 | 4.9 |
| Slope-Hunter | -0.026 | 0.032 | 61.0 | -0.069 | 0.035 | 69.6 | -0.256 | 0.037 | 82.9 | -0.292 | 0.029 | 84.0 |
| MVIVW (including disease) | -0.372 | 0.042 | 99.8 | -0.345 | 0.058 | 100.0 | -0.331 | 0.069 | 100.0 | -0.309 | 0.071 | 100.0 |
| CWBLS (disease) | -0.377 | 0.303 | 32.3 | -0.348 | 0.061 | 96.8 | -0.334 | 0.042 | 100.0 | -0.312 | 0.031 | 100.0 |

**Supplementary Table S7:** Summary of genome-wide association studies used as data sources for the summarized data analyses performed.

| Trait | Sample size | Cases | Controls | Ancestry | Reference |
| --- | --- | --- | --- | --- | --- |
| COVID-19 (COVID versus population) | 2 942 817 | 159 840 | 2 782 977 | Multi-ancestry | CHGI, 2021 |
| COVID-19 (severe COVID versus population) | 2 401 372 | 44 986 | 2 356 386 | Multi-ancestry | CHGI, 2021 |
| COVID-19 (hospitalized versus non-hospitalized) | 87 833 | 16 512 | 71 321 | Multi-ancestry | CHGI, 2021 |
| Body mass index | 694 649 | – | – | European | Pulit et al, 2019 |
| C-reactive protein | 575 531 | – | – | European | Said et al, 2022 |
| Lymphocyte count | 746 667 | – | – | European | Chen et al, 2020 |
| Type 1 diabetes | 520 580 | 18 942 | 501 638 | Multi-ancestry | Chiou et al, 2021 |
| Diabetic kidney disease | 19 300 | 7 247 | 12 053 | European | Salem et al, 2019 |
| Hypothyroidism | 1 432 752 | 155 096 | 1 277 656 | Multi-ancestry | Kurki et al, 2023 |

## References

[1] Davey Smith G, Ebrahim S. ‘Mendelian randomization’: can genetic epidemiology contribute to understanding environmental determinants of disease? International Journal of Epidemiology 2003; 32(1):1–22, doi:10.1093/ije/dyg070.

[2] Burgess S, Thompson SG. Mendelian randomization: methods for causal inference using genetic variants. Chapman & Hall, Boca Raton, FL, 2021.

[3] Lawlor D, Harbord R, Sterne J, Timpson N, Davey Smith G. Mendelian randomization: using genes as instruments for making causal inferences in epidemiology. Statistics in Medicine 2008; 27(8):1133–1163, doi:10.1002/sim.3034.

[4] Greenland S. An introduction to instrumental variables for epidemiologists. International Journal of Epidemiology 2000; 29(4):722–729, doi:10.1093/ije/29.4.722.

[5] Davies NM, Holmes MV, Davey Smith G. Reading Mendelian randomisation studies: a guide, glossary, and checklist for clinicians. British Medical Journal 2018; 362:k601, doi: 10.1136/bmj.k601.

[6] Thanassoulis G, O’Donnell C. Mendelian randomization: nature’s randomized trial in the post-genome era. Journal of the American Medical Association 2009; 301(22):2386–2388, doi:10.1001/jama.2009.812.

[7] Burgess S, Swanson SA, Labrecque JA. Are Mendelian randomization investigations immune from bias due to reverse causation? European Journal of Epidemiology 2021; 36(3):253–257, doi:10.1007/s10654-021-00726-8.

[8] Verbanck M, Chen CY, Neale B, Do R. Detection of widespread horizontal pleiotropy in causal relationships inferred from Mendelian randomization between complex traits and diseases. Nature Genetics 2018; 50(5):693–698, doi:10.1038/s41588-018-0099-7.

[9] Brumpton B, Sanderson E, Heilbron K, Hartwig FP, Harrison S, Vie GA, Cho Y, Howe LD, Hughes A, Boomsma DI, et al. Avoiding dynastic, assortative mating, and population stratification biases in Mendelian randomization through within-family analyses. Nature Communications 2020; 11:3519, doi:10.1038/s41467-020-17117-4.

[10] Davey Smith G, Lawlor D, Harbord R, Timpson N, Day I, Ebrahim S. Clustered environments and randomized genes: a fundamental distinction between conventional and genetic epidemiology. PLoS Medicine 2007; 4(12):e352, doi:10.1371/journal.pmed.0040352.

[11] Taylor M, Tansey KE, Lawlor DA, Bowden J, Evans DM, Davey-Smith G, Timpson NJ. Testing the principles of Mendelian randomization: Opportunities and complications on a genomewide scale. bioRxiv 2017; 124362, doi:10.1101/124362.

[12] Ferolito BR, Dashti H, Giambartolomei C, Peloso GM, Golden DJ, Gravel-Pucillo K, Rasooly D, Horimoto AR, Matty R, Gaziano L, et al. Leveraging large-scale biobanks for therapeutic target discovery. medRxiv 2025; :2025.02.10.25321 487 doi:10.1101/2025.02.10.25321487.

[13] Didelez V, Meng S, Sheehan N. Assumptions of IV methods for observational epidemiology. Statistical Science 2010; 25(1):22–40, doi:10.1214/09-sts316.

[14] Swanson SA, Tiemeier H, Ikram MA, Hernán MA. Nature as a trialist?: Deconstructing the analogy between Mendelian randomization and randomized trials. Epidemiology 2017; 28(5):653–659, doi:10.1097/ede.0000000000000699.

[15] Hughes RA, Davies NM, Davey Smith G, Tilling K. Selection bias when estimating average treatment effects using one-sample instrumental variable analysis. Epidemiology 2019; 30(3):350–357.

[16] Gkatzionis A, Burgess S. Contextualizing selection bias in Mendelian randomization: how bad is it likely to be? International Journal of Epidemiology 2019; 48(3):691–701, doi: 10.1093/ije/dyy202.

[17] Munafó MR, Tilling K, Taylor AE, Evans DM, Davey Smith G. Collider scope: when selection bias can substantially influence observed associations. International Journal of Epidemiology 2018; 47(1):226–235.

[18] VanderWeele TJ, Knol MJ. Interpretation of subgroup analyses in randomized trials: heterogeneity versus secondary interventions. Annals of Internal Medicine 2011; 154(10):680–683.

[19] Dudbridge F, Allen RJ, Sheehan NA, Schmidt AF, Lee JC, Jenkins RG, Wain LV, Hingorani AD, Patel RS. Adjustment for index event bias in genome-wide association studies of subsequent events. Nature Communications 2019; 10(1):1561.

[20] Canan C, Lesko C, Lau B. Instrumental variable analyses and selection bias. Epidemiology 2017; 28(3):396–398.

[21] Heckman JJ. Sample selection bias as a specification error. Econometrica 1979; 47(1):153– 161.

[22] Mahmoud O, Dudbridge F, Davey Smith G, Munafo M, Tilling K. A robust method for collider bias correction in conditional genome-wide association studies. Nature Communications 2022; 13(1):619.

[23] Burgess S, Thompson SG. Multivariable Mendelian randomization: the use of pleiotropic genetic variants to estimate causal effects. American Journal of Epidemiology 2015; 181(4):251–260, doi:10.1093/aje/kwu283.

[24] Cai S, Dudbridge F. Estimating causal effects on a disease progression trait using bivariate Mendelian randomisation. Genetic Epidemiology 2025; 49(1):e22.600.

[25] Gill D, Georgakis MK, Walker VM, Schmidt AF, Gkatzionis A, Freitag DF, Finan C, Hingorani AD, Howson JM, Burgess S, et al. Mendelian randomization for studying the effects of perturbing drug targets. Wellcome Open Research 2021; 6:16, doi:10.12688/wellcomeopenres.16544.2.

[26] Burgess S, Mason AM, Grant AJ, Slob EA, Gkatzionis A, Zuber V, Patel A, Tian H, Liu C, Haynes WG, et al. Using genetic association data to guide drug discovery and development: Review of methods and applications. The American Journal of Human Genetics 2023; 110(2):195–214.

[27] Schmidt AF, Finan C, Gordillo-Marañón M, Asselbergs FW, Freitag DF, Patel RS, Tyl B, Chopade S, Faraway R, Zwierzyna M, et al. Genetic drug target validation using Mendelian randomisation. Nature Communications 2020; 11:3255, doi:10.1038/s41467-020-16969-0.

[28] Karhunen V, Woolf B, Bhatnagar P, Gill D, Burgess S. Integrating genetic data with biological insight: a practical guide to cis-Mendelian randomization. American Journal of Human Genetics 2026; Under consideration (to update).

[29] Burgess S, Davey Smith G, Davies NM, Dudbridge F, Gill D, Glymour MM, Hartwig FP, Holmes MV, Minelli C, Relton CL, et al. Guidelines for performing Mendelian randomization investigations. Wellcome Open Research 2020; 4:186, doi:10.12688/wellcomeopenres.15555.1.

[30] Tchetgen Tchetgen E, Walter S, Vansteelandt S, Martinussen T, Glymour M. Instrumental variable estimation in a survival context. Epidemiology 2015; 26(3):402–410, doi:10.1097/ede.0000000000000262.

[31] Greenland S, Robins J, Pearl J. Confounding and collapsibility in causal inference. Statistical Science 1999; 14(1):29–46, doi:10.1214/ss/1009211805.

[32] Burgess S. Commentary: Consistency and collapsibility: are they crucial for instrumental variable analysis with a survival outcome in Mendelian randomization? Epidemiology 2015; 26(3):411–413.

[33] Gkatzionis A, Seaman SR, Hughes RA, Tilling K. Relationship between collider bias and interactions on the log-additive scale. Statistical Methods in Medical Research 2025; 34(6):1063–1078.

[34] Gkatzionis A, Tchetgen Tchetgen EJ, Heron J, Northstone K, Tilling K. Using instruments for selection to adjust for selection bias in Mendelian randomization. Statistics in Medicine 2024; 43(22):4250–4271.

[35] Puhani P. The Heckman correction for sample selection and its critique. Journal of Economic Surveys 2000; 14(1):53–68.

[36] Toomet O, Henningsen A. Sample selection models in R: package sampleSelection. Journal of Statistical Software 2008; 27(7):1–23.

[37] Burgess S, Butterworth AS, Thompson SG. Mendelian randomization analysis with multiple genetic variants using summarized data. Genetic Epidemiology 2013; 37(7):658– 665, doi:10.1002/gepi.21758.

[38] Burgess S, Dudbridge F, Thompson SG. Combining information on multiple instrumental variables in Mendelian randomization: comparison of allele score and summarized data methods. Statistics in Medicine 2016; 35(11):1880–1906, doi:10.1002/sim.6835.

[39] Slob E, Burgess S. A comparison of robust Mendelian randomization methods using summary data. Genetic Epidemiology 2020; 44(4):313–329, doi:10.1002/gepi.22295.

[40] Yao M, Miller GW, Vardarajan BN, Baccarelli AA, Guo Z, Liu Z. Deciphering proteins in Alzheimer’s disease: A new Mendelian randomization method integrated with AlphaFold3 for 3D structure prediction. Cell Genomics 2024; 4(12):100 700.

[41] Hartwig FP, Davey Smith G, Bowden J. Robust inference in summary data Mendelian randomisation via the zero modal pleiotropy assumption. International Journal of Epidemiology 2017; 46(6):1985–1998, doi:10.1093/ije/dyx102.

[42] Cai S, Hartley A, Mahmoud O, Tilling K, Dudbridge F. Adjusting for collider bias in genetic association studies using instrumental variable methods. Genetic Epidemiology 2022; 46(5-6):303–316.

[43] Burgess S, Thompson SG. Bias in causal estimates from Mendelian randomization studies with weak instruments. Statistics in Medicine 2011; 30(11):1312–1323, doi: 10.1002/sim.4197.

[44] Sanderson E, Davey Smith G, Windmeijer F, Bowden J. An examination of multivariable Mendelian randomization in the single sample and two-sample summary data settings. International Journal of Epidemiology 2019; 48(3):713–727, doi:10.1093/ije/dyy262.

[45] Patel A, Lane J, Burgess S. Weak instruments in multivariable Mendelian randomization: methods and practice. arXiv 2024; :2408.09 868v1.

[46] Ye T, Shao J, Kang H. Debiased inverse-variance weighted estimator in two-sample summary-data Mendelian randomization. The Annals of Statistics 2021; 49(4):2079–2100.

[47] Wu Y, Kang H, Ye T. A more robust approach to multivariable Mendelian randomization. Biometrika 2025; 112(4):asaf053.

[48] Donovan K, Torres J, Zhu D, Herrington WG, Staplin N. An application of the MR-Horse method to reduce selection bias in genome-wide association studies of disease progression. European Journal of Human Genetics 2025; 33:1677–1683.

[49] Grant AJ, Burgess S. A Bayesian approach to Mendelian randomization using summary statistics in the univariable and multivariable settings with correlated pleiotropy. The American Journal of Human Genetics 2024; 111(1):165–180.

[50] Sudhakar M, Winfred SB, Meiyazhagan G, Venkatachalam DP. Mechanisms contributing to adverse outcomes of COVID-19 in obesity. Molecular and Cellular Biochemistry 2022; 477(4):1155–1193.

[51] Gao M, Piernas C, Astbury NM, Hippisley-Cox J, O’Rahilly S, Aveyard P, Jebb SA. Associations between body-mass index and COVID-19 severity in 6.9 million people in England: a prospective, community-based, cohort study. The Lancet Diabetes & Endocrinology 2021; 9(6):350–359.

[52] Ponsford MJ, Gkatzionis A, Walker VM, Grant AJ, Wootton RE, Moore LS, Fatumo S, Mason AM, Zuber V, Willer C, et al. Cardiometabolic traits, sepsis and severe COVID-19: a Mendelian randomization investigation. Circulation 2020; 142:1791–1793, doi:10.1161/CIRCULATIONAHA.120.050753.

[53] Group RC. Tocilizumab in patients admitted to hospital with COVID-19 (RECOVERY): a randomised, controlled, open-label, platform trial. Lancet 2021; 397(10285):1637–1645.

[54] COVID-19 Host Genetics Initiative. Mapping the human genetic architecture of COVID-19. Nature 2021; 600(7889):472–477.

[55] Pulit SL, Stoneman C, Morris AP, Wood AR, Glastonbury CA, Tyrrell J, Yengo L, Ferreira T, Marouli E, Ji Y, et al. Meta-analysis of genome-wide association studies for body fat distribution in 694 649 individuals of European ancestry. Human Molecular Genetics 2019; 28(1):166–174, doi:10.1093/hmg/ddy327.

[56] Daghlas I, Gill D. Mendelian randomization as a tool to inform drug development using human genetics. Cambridge Prisms: Precision Medicine 2023; 1:e16.

[57] Said S, Pazoki R, Karhunen V, Võsa U, Ligthart S, Bodinier B, Koskeridis F, Welsh P, Alizadeh BZ, Chasman DI, et al. Genetic analysis of over half a million people characterises C-reactive protein loci. Nature communications 2022; 13:2198.

[58] Sudlow C, Gallacher J, Allen N, Beral V, Burton P, Danesh J, Downey P, Elliott P, Green J, Landray M, et al. UK Biobank: an open access resource for identifying the causes of a wide range of complex diseases of middle and old age. PLOS Medicine 2015; 12(3):e1001.779.

[59] Hu S, Ferreira LA, Shi S, Hellenthal G, Marchini J, Lawson DJ, Myers SR. Fine-scale population structure and widespread conservation of genetic effect sizes between human groups across traits. Nature Genetics 2025; 57(2):379–389.

[60] Hamilton F, Schurz H, Yates TA, Gilchrist JJ, Möller M, Naranbhai V, Ghazal P, Timpson NJ, Akhtar S, Anwar M, et al. Altered IL-6 signalling and risk of tuberculosis: a multi-ancestry mendelian randomisation study. The Lancet Microbe 2025; 6(1):100 922.

[61] Butler-Laporte G, Nakanishi T, Mooser V, Morrison DR, Abdullah T, Adeleye O, Mamlouk N, Kimchi N, Afrasiabi Z, Rezk N, et al. Vitamin D and Covid-19 susceptibility and severity: a Mendelian randomization study (version 2). medRxiv 2020; :2020.09.08.20190 975 URL https://www.medrxiv.org/content/10.1101/2020.09.08.20190975v2.

[62] Guo Q, Burgess S, Turman C, Bolla MK, Wang Q, Lush M, Abraham J, Aittomäki K, Andrulis IL, Apicella C, et al. Body mass index and breast cancer survival: a Mendelian randomization analysis. International Journal of Epidemiology 2017; 46(6):1814–1822, doi:10.1093/ije/dyx131.

[63] Grant AJ, Burgess S. Pleiotropy robust methods for multivariable Mendelian randomization. Statistics in Medicine 2021; 40(26):5813–5830, doi:10.1002/sim.9156.

[64] Lawton M, Ben-Shlomo Y, Gkatzionis A, Hu MT, Grosset D, Tilling K. Two sample Mendelian Randomisation using an outcome from a multilevel model of disease progression. European Journal of Epidemiology 2024; 39(5):521–533.

